# Climate, demography, immunology, and virology combine to drive two decades of dengue virus dynamics in Cambodia

**DOI:** 10.1101/2022.06.08.22276171

**Authors:** Cara E. Brook, Carly Rozins, Jennifer A. Bohl, Vida Ahyong, Sophana Chea, Liz Fahsbender, Rekol Huy, Sreyngim Lay, Rithea Leang, Yimei Li, Chanthap Lon, Somnang Man, Mengheng Oum, Graham R. Northrup, Fabiano Oliveira, Andrea R. Pacheco, Daniel M. Parker, Katherine Young, Michael Boots, Cristina M. Tato, Joseph L. DeRisi, Christina Yek, Jessica E. Manning

**Affiliations:** Department of Ecology and Evolution, University of Chicago, Chicago, Illinois, USA; Department of Science and Technology Studies, York University, Toronto, Canada; Laboratory of Malaria and Vector Research, National Institute of Allergy and Infectious Diseases, National Institutes of Health, Rockville, Maryland, USA; Chan Zuckerberg Biohub, San Francisco, California, USA; International Center of Excellence in Research, National Institute of Allergy and Infectious Diseases, National Institutes of Health, Phnom Penh, Cambodia; Chan Zuckerberg Initiative, San Francisco, California, USA; National Center for Parasitology, Entomology, and Malaria Control, Phnom Penh, Cambodia; Center for Computational Biology, University of California, Berkeley, California, USA; Department of Population Health and Disease Prevention, University of California, Irvine, California, USA; Department of Epidemiology and Biostatistics, University of California, Irvine, California, USA; Department of Biological Sciences, University of Texas, El Paso, Texas, USA; Department of Integrative Biology, University of California, Berkeley, California, USA

**Keywords:** dengue, genomic epidemiology, force of infection, wavelet decomposition

## Abstract

The incidence of dengue virus disease has increased globally across the past half-century, with highest number of cases ever reported in 2019. We analyzed climatological, epidemiological, and phylogenomic data to investigate drivers of two decades of dengue in Cambodia, an understudied endemic setting. Using epidemiological models fit to a 19-year dataset, we first demonstrate that climate-driven transmission alone is insufficient to explain three epidemics across the time series. We then use wavelet decomposition to highlight enhanced annual and multiannual synchronicity in dengue cycles between provinces in epidemic years, suggesting a role for climate in homogenizing dynamics across space and time. Assuming reported cases correspond to symptomatic secondary infections, we next use an age-structured catalytic model to estimate a declining force of infection for dengue through time, which elevates the mean age of reported cases in Cambodia. Reported cases in >70 year-old individuals in the most recent 2019 epidemic are best explained when also allowing for waning multitypic immunity and repeat symptomatic infections in older patients. We support this work with phylogenetic analysis of 192 dengue virus (DENV) genomes that we sequenced between 2019-2022, which document emergence of DENV-2 Cosmopolitan Genotype-II into Cambodia. This lineage demonstrates phylogenetic homogeneity across wide geographic areas, consistent with invasion behavior and in contrast to high phylogenetic diversity exhibited by endemic DENV-1. Finally, we simulate an age-structured, mechanistic model of dengue dynamics to demonstrate how expansion of an antigenically distinct lineage that evades preexisting multitypic immunity effectively reproduces the older-age infections witnessed in our data.

**CLINICAL TRIAL NUMBERS:** NCT04034264 and NCT03534245.

**SIGNIFICANCE STATEMENT:** The year 2019 witnessed the highest number of dengue cases ever reported, including in Cambodia, a Southeast Asian country with endemic transmission. We analyzed 19 years of national dengue surveillance data for Cambodia to demonstrate how increasing temperature and precipitation enhance similarity in dengue incidence across space and time, particularly in epidemic years. We document how two decades of demographic transition has depressed the rate at which dengue infections are acquired, thus increasing the age of reported infection. In 2019, expansion of a genetically distinct DENV-2 lineage into Cambodia likely underpinned repeated symptomatic infections in older-age individuals to drive high caseloads. As climates warm, we anticipate more synchronized dynamics globally and a shifting burden of symptomatic disease into older cohorts.

## INTRODUCTION

Dengue virus (DENV) transmission has increased dramatically over the past two decades, culminating in 2019 with the highest number of global cases ever reported (>5.2 million) to the World Health Organization (WHO) (1). Since nearly three-quarters of DENV infections are estimated to be clinically inapparent (thereby unreported), these counts represent a vast underestimate of the true scale of dengue burden (2). DENV is a flavivirus primarily transmitted by the *Aedes aegypti* mosquito, an ubiquitous arthropod vector in tropical and subtropical regions (2). DENV is comprised of four antigenically distinct serotypes—each of which is further subdivided among four to seven distinct genotypes; infection with one serotype is thought to result in lifelong immunity to that same serotype (homotypic immunity) but only temporary (up to two-year) protection against different serotypes (heterotypic immunity) (3). Heterotypic secondary infections are often more clinically severe due to an interaction with pre-existing flavivirus-specific antibodies known as antibody-dependent enhancement (ADE) (4, 5). Southeast Asia (SEA) reports ∼70% of dengue cases globally (1). Historically, dengue has been classified as an urban disease, though recent studies highlighting the importance of microscale transmission in generating DENV diversity in urban Bangkok predict that DENV transmission will intensify across SEA as peri-urban settings become better connected (6–8). Cambodia, at a population size of 16 million, is currently the least urbanized country in SEA (9) but exhibits significant peri-urban sprawl surrounding major metropolitan centers like the capital city, Phnom Penh (6, 7, 9). Urbanization and climatic changes are increasing the population at-risk for dengue in Cambodia and elsewhere (10–12).

Dengue was first detected in Cambodia in 1963 (13), though political instability and civil war in the late 1970s delayed formalization of a passive surveillance policy until 1980 (14). Once adopted, this policy required reporting of clinically diagnosed cases from public health centers and hospitals to the national level. In 2001, the National Dengue Control Program (NDCP) was inaugurated in Cambodia, and in 2002, the NDCP formally implemented the WHO clinical case definitions of dengue and its complications as criteria for the surveillance program (14, 15). The NDCP surveillance system is largely limited to clinicosyndromic diagnoses, meaning that documented cases often correspond to more severe heterotypic secondary DENV infections (16), which are reported collectively without the ability to systematically distinguish serotypes (15). Nonetheless, the NDCP also instituted some active surveillance efforts in Cambodia in 2001, which included limited molecular testing to identify distinct serotypes at sentinel sites in four provinces, which were expanded to 15 provinces by 2021 (14, 15).

Over the past two decades, all four dengue serotypes have been detected in Cambodia by molecular surveillance (15), though cases have been largely dominated by one or two serotypes in a given year—with DENV-1 and DENV-3 most common in the early 2000s and DENV-1 and DENV-2 predominant in the last decade (15). Cambodia witnessed three major dengue epidemics across this period–in 2007, 2012, and 2019 (15). The first epidemic, in 2007, occurred coincidentally with a genotype replacement event in DENV-1 (17, 18), though cases were dominated by DENV-3, marking the last year of this serotype’s dominance in the region (14, 18). The second epidemic, in 2012, has been largely attributed to DENV-1 (15, 19–21), and the third, in 2019, appears to have been driven by co-circulation of both DENV-1 and DENV-2 (15). Consistent with the global trends (1), Cambodia suffered its worst dengue epidemic on record in 2019, with approximately 40,000 total cases reported across all 25 provinces—a likely drastic underestimation of the true disease burden (22). The past twenty years of surveillance in Cambodia have witnessed, on average, steadily increasing dengue incidence, coupled with a steady increase in the mean age of reported dengue infection (15). The increased age of reported infection has been anecdotally attributed to an aging population following demographic transition, as similar phenomena have been documented previously in neighboring Thailand (23, 24) and in Nicaragua (25).

Explosive periodic outbreaks are a hallmark of dengue virus disease, though the multiple drivers of these phenomena have long been debated (24, 26–29). Seasonal climate cycles are a strong predictor of annual cycling for many arboviruses, including dengue (10, 11), and climate has been implicated as a possible driver of multiannual dengue periodicity, as well. In Thailand, multiannual dengue cycles demonstrate coherence with El Niño phenomena (29), and epidemic years exhibit more synchronized dynamics across latitudes (29), as well as higher correlation with local temperature than do inter-epidemic periods (28). The interaction of demography and heterotypic immunity is also thought to play a role in driving multiannual dengue cycles (24, 26, 30), which, in Thailand, show elongated periodicity as a result of declining birth rates and slower build-up of the susceptible population over the past half-century (24). Virology also contributes to dengue epidemics, which have, historically, been linked to turnover in the dominant regional serotype (31–33) or to replacement of a dominant viral genotype within a single serotype (18, 34). Genotype-specific antigenic diversity is increasingly recognized within traditional serotype classifications for dengue (35), and recent work links the magnitude of periodic dengue epidemics to this antigenic evolution (34, 35). Large epidemics tend to result from the takeover of DENV lineages most antigenically distinct from previously circulating strains of the same serotype or most antigenically similar to lineages of a different serotype (34).

Here, we explore the dynamics of dengue virus transmission across the past two decades in peri-urban Cambodia, investigating the potential mechanisms that underly periodic epidemics, particularly the epidemic of 2019. We queried a 19-year dataset (2002–2020) of serotype-agnostic dengue case counts from the NDCP, aggregated at the province level, to, first, interrogate the role that climate played in driving epidemics in 2007, 2012, and 2019, and second, more generally, explore the impact of climate on annual and multiannual cycling across the time series. We, third, fit catalytic models to the age-structured incidence of reported dengue disease to estimate the annual force of infection (FOI), at the province level, across this time series (24, 36, 37), then considered the national data in the context of our own active febrile surveillance study carried out in peri-urban Kampong Speu and surrounding provinces from 2019-2022 (38). Whole genome sequencing of DENV from serum samples collected in our active surveillance program, combined with additional sequences from NDCP surveillance, identified the first record of the DENV-2 Cosmopolitan lineage (Genotype-II (39)) ever documented in Cambodia (40). Phylodynamic analysis of the spread of the DENV-2 Cosmopolitan lineage in this region suggests that this genotype caused pathogenic infections in older age individuals, contributing to the largest documented dengue outbreak on record for Cambodia in 2019. Finally, we constructed an age-structured discrete-time, dynamical model to simulate the interplay of climate, demography, immunology, and virology which combine to structure two decades of dengue dynamics in Cambodia.

## RESULTS

### Though warming temperatures correlate with increasing dengue incidence in Cambodia, epidemic years are not consistent climate anomalies

Because of the widely acknowledged role of climate as a driver of arboviral disease globally (10, 11), coupled with recent work from Thailand that highlights coherence between El Niño climate anomalies and dengue epidemics (29), we first explored the influence of changing temperature and precipitation on dengue caseload in Cambodia. To this end, we aggregated high-resolution temperature and precipitation data at the Cambodian province level, across two-week intervals from 2002-2019 and sought to address the extent to which epidemic years represented climatic anomalies over this time period; we defined ‘anomalies’ as any years in which temperature or precipitation records demonstrated significant deviations from average trends through time, as calculated using generalized additive models (GAMs) (41). Across the time series, temperature peaked in the first half of each year, between April and July, preceding the peak in dengue caseload (Fig. S1). Precipitation peaked in the latter half of each year, between August and October (Fig. S2), preceding the onset of the following year’s dengue season. GAMs (41) demonstrated that, after controlling for intra-annual variation, temperature has increased significantly across the past two decades in all provinces; no significant interannual changes were detected for precipitation (Fig. S3-S4; Table S1). Additional GAMs and climate data normalized into z-scores indicated that the epidemic years of 2012 and 2019 were anomalously warm, while 2007 was anomalously cool, consistent with the observed interannual increase in temperature (Fig. S5). Years 2015-2016, which spanned a major El Nino event in SEA (but did not correspond to a dengue epidemic in Cambodia) (42), were also significantly warmer than average. By contrast, precipitation showed no anomalies in the year preceding each of the three major dengue epidemics (Fig. S6). We concluded that, while both temperature and dengue incidence increased across our time series, epidemic years could not be consistently distinguished by any consistently aberrant climatic profile.

### Climate-informed transmission rates failed to recover epidemic dynamics in a TSIR model

To formally interrogate the role of climate as a driver of dengue epidemics in Cambodia, we developed a simple, climate-informed time series Susceptible-Infected-Recovered (TSIR) model (43–45), which we fit to dengue case counts, aggregated over two week intervals at the province level, for the three inter-epidemic periods (2002-2006, 2008-2011, and 2013-2018). Typically used to describe the transmission dynamics of perfectly immunizing childhood infections, such as measles (44, 45), TSIR has been applied to dengue dynamics previously and offers an effective means by which to isolate the impact of climate on transmission, despite oversimplifying the multiple serotype dynamics of dengue virus disease (46–50).

We used biweekly transmission rates recovered from TSIR fits to the inter-epidemic periods, as well as climate-informed transmission rates for epidemic years projected from province-specific lagged temperature and precipitation data, to predict epidemic-year cases (Fig. 1; *Methods; SI Appendix*). We found that TSIR successfully recaptured the timing of annual dengue epidemics across the 22 provinces considered, with the recovered transmission rate peaking between May and August, slightly preceding reported cases. The magnitude and timing of transmission varied by province and between the three inter-epidemic periods, showing no consistent pattern of directional change in magnitude with time (Table S2). TSIR-estimated transmission was significantly positively associated with higher temperature and precipitation (lagged such that climate variables preceded transmission by a median 3.5 months for temperature and 1 month for precipitation) in the corresponding province across all inter-epidemic periods (Table S3-S4). More rapid transmission gains were observed for corresponding increases in temperature vs. precipitation (Fig. S7-S9; Table S4). Nonetheless, climate-informed transmission rates for epidemic years were not substantially different from rates fitted to inter-epidemic periods, reflecting the absence of major climate anomalies across the time series (Fig. S10).

**Figure 1.**
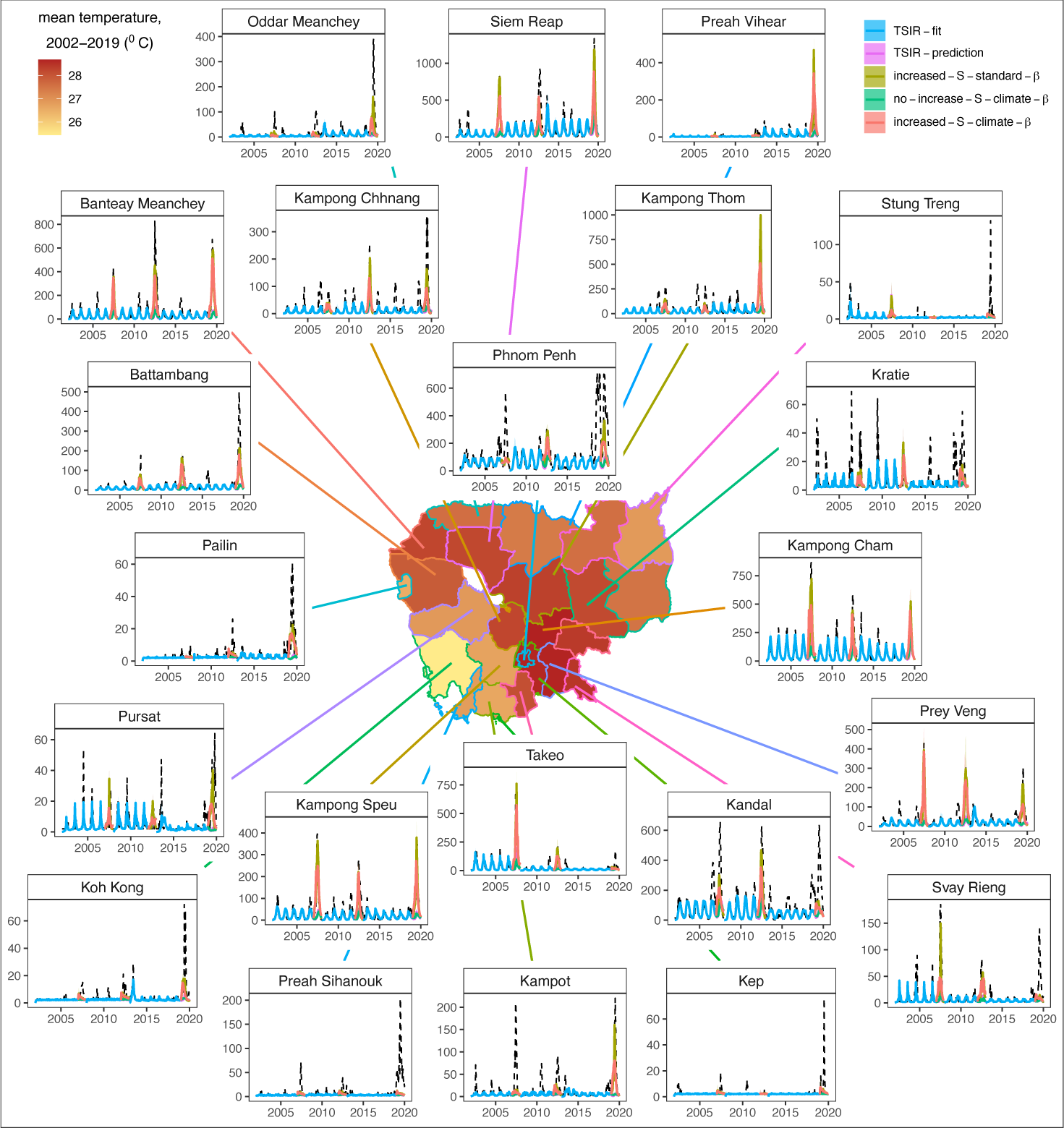
Climate-informed TSIR insights into epidemic dynamics of DENV in Cambodia. Inset panels show province level clinicosyndromic reported DENV cases (*dashed black lines*) with fitted TSIR output to the three inter-epidemic periods (2002-2006, 2008-2011, 2013-2018; *blue lines*) and TSIR projections for epidemic years (2007, 2012, 2019) under trained biweekly transmission rate (𝛽) estimates (*pink lines*), incorporating a factorial increase in the susceptible population (*gold lines*), using a climate-projected 𝛽 estimated from lagged temperature and precipitation data by province (*green lines*), or using both climate-projected 𝛽 and a factorial increase in the susceptible population (*red lines*) (*SI Appendix,* Table S2, S5). The center map shows province level administrative boundaries for Cambodia, shaded by the mean biweekly temperature in 2020.

As expected, TSIR underpredicted the magnitude of epidemic year caseloads for all provinces; climate-informed transmission improved TSIR projections but still failed to recover epidemic peaks (Fig. 1). Experimentally, we amplified the susceptible population input at the beginning of each epidemic year in our model, which facilitated recovery of DENV caseloads (*Methods*) (50). In the absence of climate input into the transmission rate, the proportional increase in susceptible population at the beginning of each epidemic year needed to recover the epidemic peak was a median 1.8x for 2007, 2.10x for 2012, and 2.17x for 2019 (Table S5). The proportional increase in the susceptible population needed to recover the epidemic peak was not significantly lower when epidemic year cases were projected using climate-informed transmission rates for 2007 and 2012 (paired student’s t-test: [2007] t=0.33, p>0.1; [2012] t=-0.52;p>0.1) and only marginally lower for 2019 (t=1.31, p=0.1), suggesting (at most) a minimal role for temperature and precipitation in driving epidemic dynamics (Table S5). For many provinces, the combination of climate-informed transmission rate with susceptible augmentation still fell short of effectively capturing epidemic caseloads, highlighting the need for alternative explanations for these high transmission years.

### Wavelet analysis showed enhanced synchrony in dengue dynamics across provinces and climate time series in epidemic years

Though TSIR modeling failed to recover a clear role for climate in driving major dengue epidemics in Cambodia, the analysis did highlight a critical influence of climate on the intraannual timing of cases and the interannual periodicity of dengue dynamics. Prior work in Thailand has demonstrated enhanced synchronicity between dengue incidence and climate, and between geographically distant localities, during major epidemics (28, 29), as well as a link between multiannual climate cycles and higher burden epidemics (29). We hypothesized that these multiannual climate cycles not considered in TSIR might also be important in the Cambodia system. To this end, we undertook wavelet decomposition of the biweekly dengue time series and the corresponding climate data, both at the province level, to extract and disaggregate annual and multiannual trends. We found elevated amplitude and statistically significant peaks in average wavelet power of both annual (Fig. 2A, S11A) and multiannual (Fig. 2B, S11B) cycles in dengue incidence during the three epidemic years (51). Elevated amplitudes for multiannual cycles were also observed during the 2015-2016 El Niño event in more northerly provinces (Fig. 2B), including those abutting Thailand, where associations between dengue and El Niño have been previously established (29). We additionally observed significantly elevated synchronicity between provinces in the timing and amplitude of yearly dengue incidence across epidemic years, as measured both by pairwise Pearson’s correlation coefficient (Fig. 2C) and cross-wavelet power (Fig. S11). Synchronicity in the annual incidence data was negatively associated with geographic distance between provinces, though patterns were less clear than have been described elsewhere (29) (Fig. S12). High synchronicity between annual case data for paired provinces was significantly positively associated with high temperature, precipitation, and population size of a focal province (Fig. S12; Table S6).

**Figure 2.**
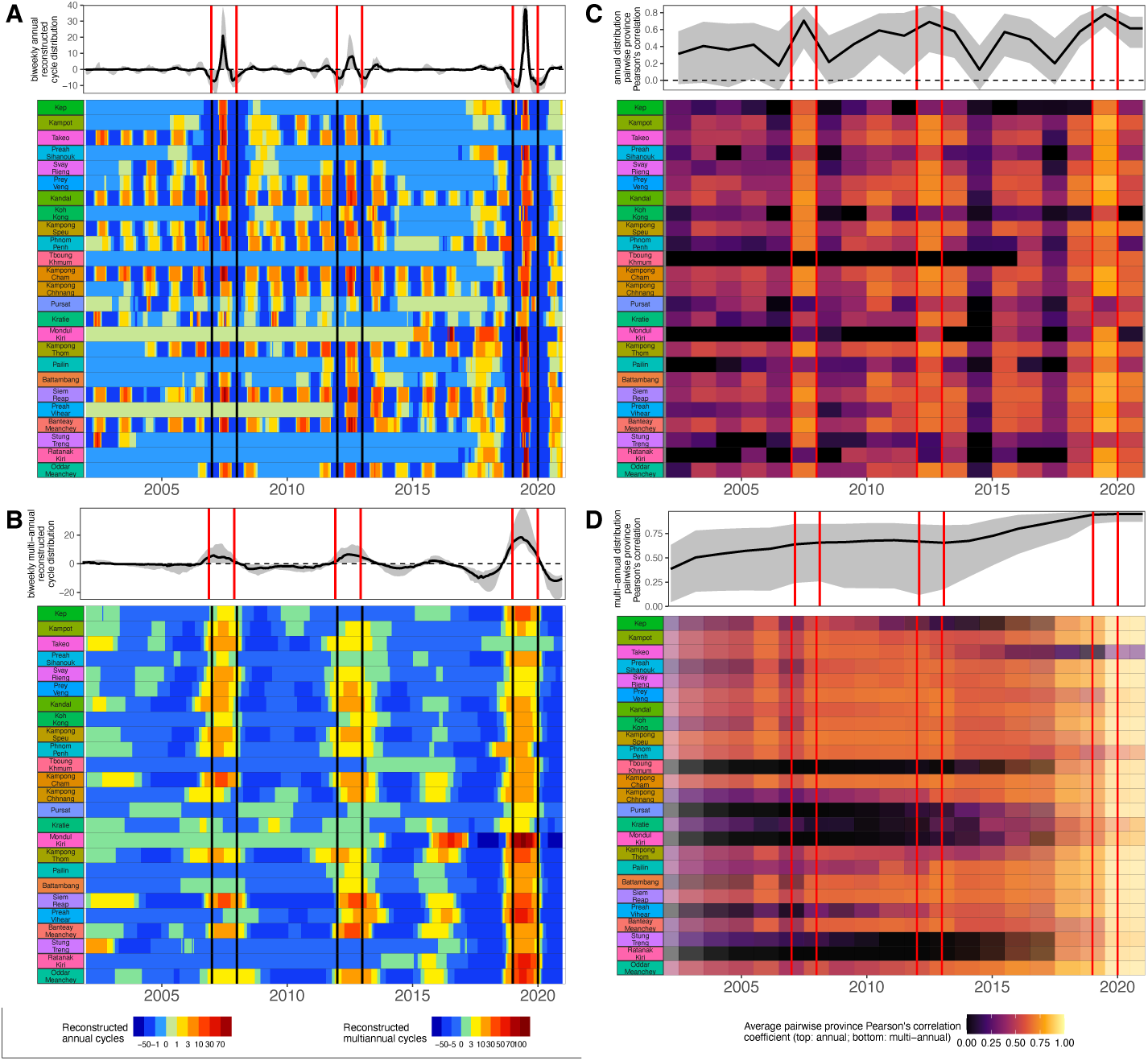
Wavelet reconstructions show heightened synchrony in epidemic years. Reconstructed **A** annual and **B** multiannual dengue cycles by province, by year from NDCP incidence per 100,000 population. **C** Mean pairwise Pearson’s correlation coefficient (𝜌) for annual dengue incidence between focal province and all other provinces through time. **D** Mean 𝜌 comparing province-to-province reconstructed multiannual dengue cycles across a 5-year sliding window, with overlapping window frames plotted (partially translucent) atop one another. In all panels, epidemic years are highlighted by vertical red or black bars. Top panels indicate the distribution of corresponding values (median = solid line; max-to-min range = gray shading) observed across all provinces within each timestep. X-axis labels are marked on January 1 of the corresponding year.

For multiannual cycles, we observed a trend of steadily increasing synchronicity (Fig. 2D) and cross-wavelet power (Fig. S11D) between provinces through time, though no peaks occurred in epidemic years. Cross-wavelet power between raw dengue incidence and the corresponding, province-level mean temperature and total precipitation (grouped biweekly) also peaked in epidemic years (Fig. S13 AB). At the multiannual scale, cross-wavelet power between province-level reconstructed dengue cycles and temperature and precipitation largely increased across the time series (Fig. S13CD), though values were highest slightly preceding the 2019 epidemic and overlapping the 2015-2016 El Niño event (42). The same pattern was observed when comparing monthly reconstructed multiannual dengue cycles and the Oceanic Niño Incidence (ONI) (Fig. S13E). For temperature, precipitation, and ONI, cross-wavelet power with multiannual dengue cycles peaked earlier in the time series in more southern provinces and moved gradually northward (Fig. S13C-E). All told, wavelet analyses suggested a role for climate in synchronizing annual dengue epidemics across provinces, but only minimal support for a hypothesis of multiannual climate cycles driving periodic epidemics across our time series. In contrast to reports from two decades prior in Thailand (24), the periodicity of multiannual dengue cycles across provinces did not increase in duration throughout our time series, despite decreasing birth rates (Fig. S14). This perhaps reflects the relative slowdown in both birth and death rate declines in Cambodia over the past two decades, when compared with the dramatic declines witnessed twenty years prior (Fig. S14).

### Mean age of reported dengue infection increased across the study period, corresponding to a declining force of infection

The muted effects of climate on our epidemic time series led us to next investigate demographic and immunological drivers of dengue dynamics in Cambodia. Previous work has documented a trend of increasing mean age of reported dengue infection across the past two decades in Cambodia at the national level (15); we confirmed this to be consistent at the province level (Fig. 3A,B; Fig. S15; Table S7). Nationally, the mean age of reported dengue infection increased from 6.79 years (95% CI: 5.85-7.72) in 2002 to 10.34 years (95% CI: 9.41-11.27) in 2020 (p<0.001). Increases in age of reported cases were even more dramatic in more remote provinces: in the distant northeastern province of Mondul Kiri, for example, the mean reported age of infection increased from 3.72 years (95% CI: 1.97-5.47) in 2002 to 21.18 years (95% CI: 20.84-21.53) in 2020 (Fig. 3A; Fig. S15; Table S7).

**Figure 3.**
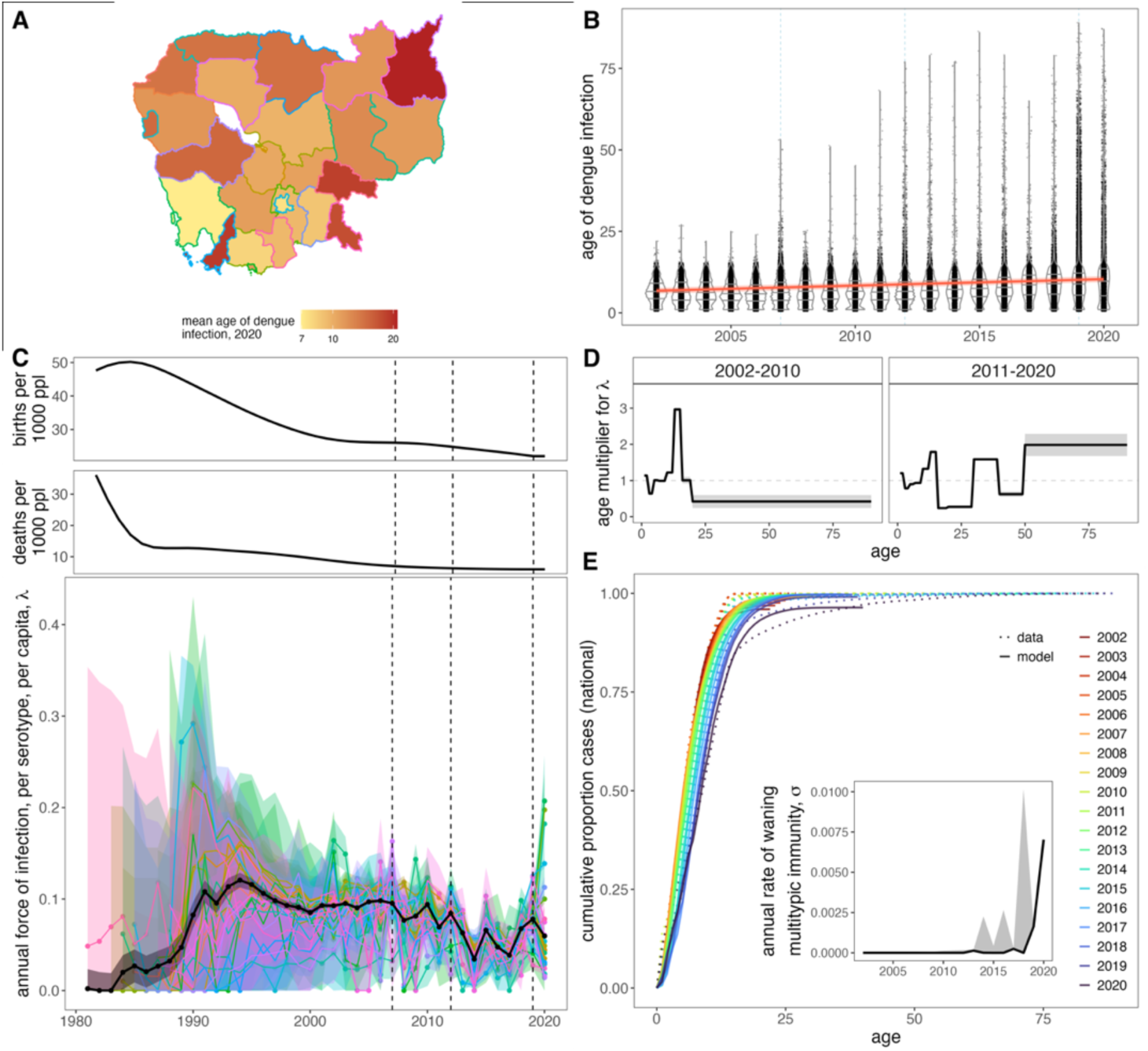
Demographic transition underpins declining force of infection and increasing age of reported dengue incidence in Cambodia. **A** Mean age of reported dengue infection, by province in the last year of the NDCP time series (2020). **B** Age distribution of reported dengue cases by year, with violin plots highlighting changes in the interquartile range of cases. The interannual trend in the mean age of dengue infection is plotted as a solid red line, with 95% confidence intervals by standard error shown as a narrow, translucent band behind it (Table S7). Epidemic years (2007, 2012, 2019) are highlighted by a light blue, dashed line in the background. **C** National (black) and province level (colored) estimates for the annual, per serotype, per capita force of infection from 1981 to 2020. 95% confidence intervals from the hessian matrix are shown as translucent shading. FOI estimates are compared against national birth and death rates for Cambodia across the time series, with epidemic years highlighted by vertical dashed lines. **D** Age modifiers to the FOI fit as shared across all provinces for 2002-2010 and 2011-2020 subsets of the data, with 95% confidence intervals from the hessian matrix are shown as translucent shading. **E** Cumulative increase in the proportion of cases reported by age at the national level, colored by year. Data are shown as dotted lines and model output as solid lines. Model includes national FOI estimates from **C**, age modification terms from **D**, and time-varying waning multitypic immunity as shown in the inset.

To interrogate the mechanistic drivers of this pattern, we estimated the annual force of infection (FOI, or 𝜆), the rate at which susceptibles become infected, by fitting a catalytic model with multiple serotype exposures to the province level data for every year in the dataset (2002–2020) and the 22 years preceding the onset of the time series, dating back to the birth year of the oldest individual in the first year of the data (Fig. 3C) (24, 36, 37). As in previous models for dengue (24, 36), we assumed that reported cases represented secondary, symptomatic infections most likely to report to public hospitals and clinics. Resulting patterns in FOI by province largely mirrored those recovered nationally (Fig. S16), demonstrating a consistently high FOI in the early 1990s, corresponding to the years in which individuals born in Cambodia’s 1980s birth pulse (which followed severe population reductions from civil war in the 1970s) would likely experience secondary infections. The FOI subsequently demonstrated a gradual decline across the time series, interspersed with local peaks in epidemic years and in 2010 and 2015.

### Age-structured FOI modifications and waning multitypic immunity in 2019-2020 improved the catalytic model’s ability to recapture observed data

We next fit 19 multiplicative age modification parameters, shared across all provinces, to allow for modulation of annual FOI across different age categories; eight age class were considered for the first half of the time series and eleven for the second, corresponding to the expanded age range of reported cases (Fig. 3D). Incorporation of age modifiers improved model fits to the data across all provinces (Table S8). Consistent with recent work in Thailand (23), we identified a high hazard of infection in adolescents (13-15 year-olds) across the time series. In the second decade, we additionally noted an elevated age-specific hazard of infection in 30-39 year-olds and >50 year-olds, corresponding to case reports in older cohorts.

Because the age range of these older cases greatly exceeded average age increases predicted by declining FOI, we reconsidered our original assumption by which reported cases corresponded to clinically apparent secondary infections only. Visualization of the age distribution of reported cases by year at both the national (Fig. 3B) and province levels (Fig. S15) indicated that, in addition to increases in the mean reported age of dengue infection, the past two decades of dengue incidence in Cambodia have also witnessed expansion in the age *range* of reported cases—such that approximately 1% of reported cases (712/68597) in 2019 occurred in individuals over the age of 45, with 61 infections reported in individuals >70 years in age. We also observed an isolated spike in the age distribution of reported cases in some provinces in the epidemic years of 2007 and 2012 (Fig. S15). To recapture these symptomatic infections in older age-individuals, we modified the multitypic catalytic modeling framework presented in previous work (24, 25, 36) to permit individuals to wane from a state of multitypic immunity back to that of monotypic immunity subject to renewed hazard of symptomatic re-infection (*SI Appendix*). Using this new modeling framework, we estimated a shared parameter across all provinces, which corresponded to the rate of waning multitypic immunity (𝜎). When we allowed 𝜎 to vary by year, we estimated a significant signature of waning multitypic immunity in the 2019 and 2020 data only (Fig. 3E, *inset*). Incorporating time-varying 𝜎 improved model fit when added both to the FOI-only model and the FOI model modified by age class (Table S8). Our best fit FOI model including both age modifications and time-varying multitypic waning immunity effectively recaptured the observed cumulative proportion of cases per age class per year at national (Fig. 3E) and province (Fig. S17) levels. In general, cases accumulated more slowly across age classes with advancing years, tracking declining FOI.

### DENV genotyping from 2019-2022 suggests a likely DENV-2 clade replacement event in Cambodia

The results of our FOI analysis led us to question whether regional turnover in the dominant DENV virus lineage could underpin the 2019 signature of immunological naivety identified in our data. Prior work emphasizes the key role that antigenic novelty, influenced by both serotype and genotype, plays in determining fitness advantages for co-circulating DENV variants (34, 35). To address this question in our system, we leveraged serum samples amassed during an active febrile surveillance study that we carried out in Kampong Speu province, adjacent to the Cambodian capital city of Phnom Penh, between July 2018 and December 2022 (38, 40). In this study, serum collected from patients reporting to the provincial referral hospital within 24 hours of fever was subject to RNA extraction and metagenomic Next Generation Sequencing (mNGS) to identify the etiology of infection (38, 40). We successfully identified serotype and corresponding genotype from mNGS data for 239 DENV-positive Kampong Speu cases. We supplemented these with samples collected from 11 cases identified in our more limited active surveillance in five neighboring provinces in 2021-2022 and 20 cases identified in routine NDCP surveillance across six provinces, also in 2021-2022 (Fig. 4A; Table S9). From this data subset of positive DENV samples, we sequenced and submitted 192 whole DENV genomes to NCBI (63 DENV-1, 120 DENV-2, and 9 DENV-4; Table S9), representing one-third of full genome DENV-1 sequences and two-thirds of full genome DENV-2 sequences currently available for Cambodia.

**Figure 4.**
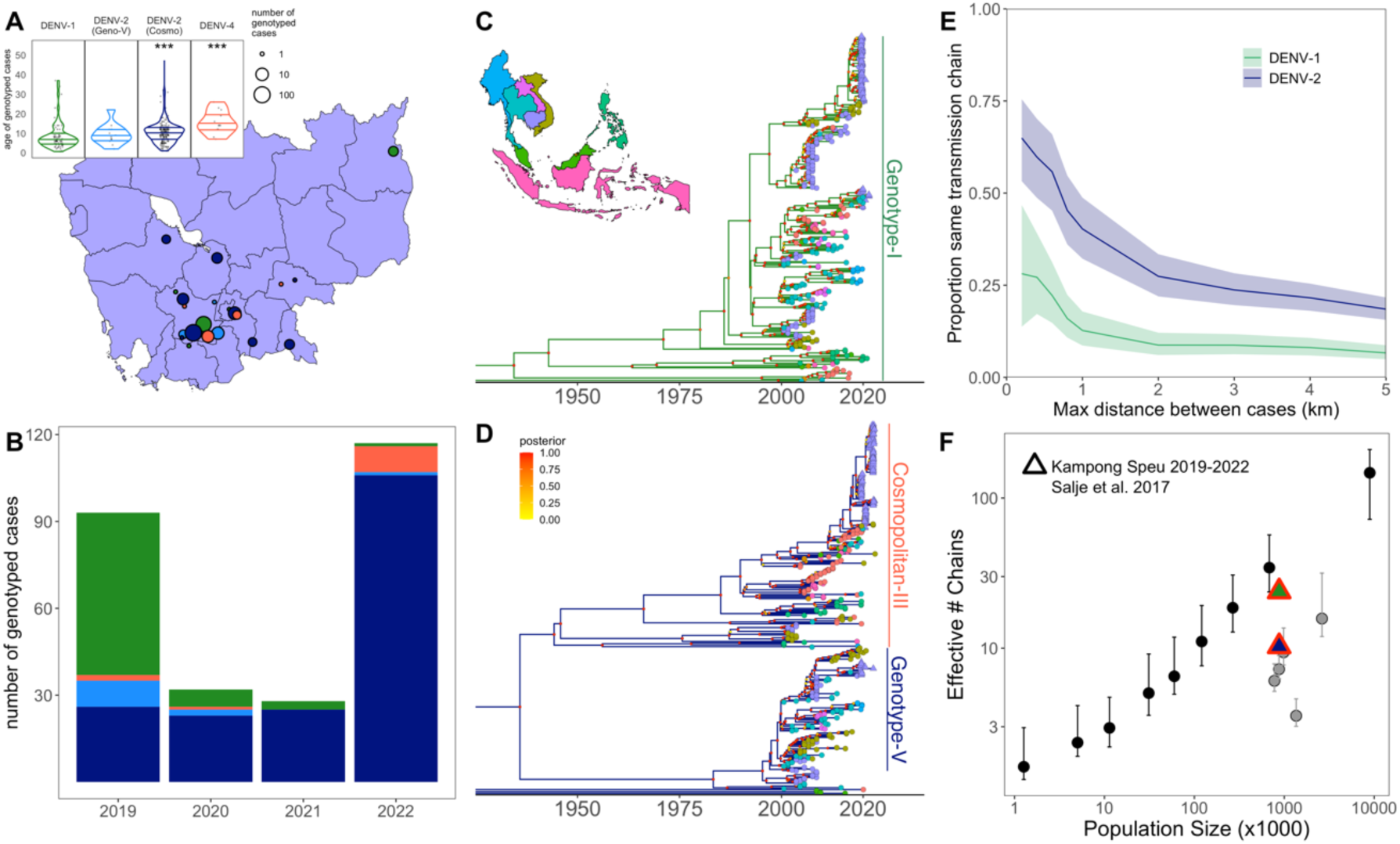
Bayesian time trees highlight geospatial structuring in evolutionary relationships for Cambodian dengue. **A** Map of Cambodia with locations of DENV serum samples genotyped from 2019-2022 in part with this study. Cases are grouped together within 10km radii. The centroid of each case cluster is plotted, with circle size corresponding to sample number and shading corresponding to serotype and genotype (DENV-1=green; DENV-2 Genotype-V = light blue; DENV-2 Cosmopolitan (Genotype-II) = navy; DENV-4=orange). The age distribution of cases by serotype and genotype is shown in the upper left; DENV-2 Cosmopolitan and DENV-4 infections occurred in significantly older age individuals than reference DENV-1 infections (linear regression; p<0.001; Table S10). **B.** Number of genotyped DENV cases by year from febrile surveillance in this study, colored by serotype and genotype as showing in panel A. **C** Map of Southeast Asia with countries colored corresponding to sequences, as shown in tip points on phylogenetic timetrees constructed using BEAST 2 for DENV-1 and **D** DENV-2. X-axis highlights divergence times between corresponding sequences. Reference sequences from GenBank are represented as circle tips and sequences contributed by active febrile surveillance in this study as triangles. Cambodia and corresponding sequences are shaded purple. Clade bars indicate the genotype of corresponding sequences within each serotype: genotype-1 for DENV-1 and Genotype-V and Cosmopolitan III (Genotype-II) for DENV-2. See Fig. S19 for a detailed inset of geographic localities for 2019-2022 Cambodia sequences. **E** Proportion of geolocated sequence pairs from panel A for DENV-1 (*green*) and DENV-2 (*blue*) genomes derived from the same transmission chain across progressively longer Euclidean distances. **F** Number of effective transmission chains for circulating DENV estimated across populations of varying densities. Black (urban) and gray (rural) circles with corresponding 95% confidence intervals depict estimates for Thailand from Salje et al. 2017 (8), while triangles depict estimates from our Kampong Speu active febrile surveillance study for DENV-1 (*green*) and DENV-2 (*blue*).

Maximum likelihood phylogenetic analysis of the resulting genomes demonstrated that most DENV-2 sequences recovered from our 2019-2022 study belonged to the DENV-2 Cosmopolitan III lineage (Genotype-II), the first record of this genotype in Cambodia, though its emergence has been reported recently in several neighboring SEA countries (Fig. S18) (52–55). All DENV-1 sequences clustered in the Genotype-1 lineage, consistent with previous genotype records in Cambodia (Fig. S18). Genotyped cases from 2019 were largely split between DENV-1 (N=56/93, 60.2%) and DENV-2 (N=35/93, 37.6%) serotypes, while those from 2020-2022 were dominated by DENV-2 (2020: N=25/32, 78.1%; 2021: 25/28, 89.3%; 2022: 107/117, 91.4%) (Fig. 4B). A small number of DENV-4 cases were also sequenced across the time series (Fig. 4B). The Cosmopolitan III lineage (Genotype-II) of DENV-2 became increasingly prevalent across the time series, as compared with the Genotype-V lineage previously recorded in Cambodia (Fig. 4B). The Cosmopolitan lineage increased in prevalence from 74.3% (26/35) of DENV-2 cases in our study in 2019 to 99.1% (106/107) of cases in 2022 (Fig. 4B). We additionally observed that the mean age of infection was significantly higher for DENV-2 Cosmopolitan cases genotyped in our study (while the mean age of DENV-2 Genotype-V infection showed no difference), as compared with reference DENV-1 case (Fig. 4A; Table S10, p<0.001). Mean age of infection was also significantly elevated for DENV-4, likely because it circulated at much lower prevalence.

We constructed serotype-specific Bayesian timetrees (56, 57) from full genome DENV sequences to assess the divergence time of 2019-2022 lineages from sequences previously reported from Cambodia and neighboring SEA countries between 2002-2022 (Fig. 4C-D; Table S9). The majority of DENV-1 lineages recovered from our study in 2019-2022 clustered closely with other DENV-1 sequences reported to NCBI from Cambodia across the past decade (Fig. 4C). By contrast, DENV-2 sequences in the Cosmopolitan III lineage (Genotype-II) demonstrated high divergence from the most recently reported Genotype-V sequences for Cambodia, with a time to most recent common ancestor (tMRCA) of approximately 77.1 years (Fig. 4D; MRC: 1945, 95% HPD: 1942-1949). Locally, within our regional data subset, sequences collected from cases coincident in geographic space and time were highly phylogenetically related (Fig. S19), supporting previous studies emphasizing the importance of microscale transmission for DENV (8). Nonetheless, we observed reduced phylogenetic diversity overall and a higher degree of phylogenetic relatedness across wider geographic distances for DENV-2 vs. DENV-1 sequences, consistent with a hypothesis of recent genotype invasion (Fig. 4E). Indeed, some 60% of paired DENV-2 sequences separated by up to 400m of geographic distance were estimated to share a MRCA within six months of the onset of the earlier case (corresponding to a shared ‘transmission chain’), as compared with only 22% of DENV-1 sequences. At a distance of up to 1 km separation, some 40% of DENV-2 pairs still belonged to the same transmission chain, in contrast to only 13% of DENV-1 sequences. Specifically for the Kampong Speu data, we additionally estimated the number of distinct DENV-2 vs. DENV-1 effective transmission chains in circulation in the region (Fig. 4F). When compared with prior work from neighboring Thailand, the number of effective transmission chains per population size for DENV-1 in Kampong Speu closely matched that predicted by population size in the endemic transmission setting of urban Bangkok (8). By contrast, the lower number of transmission chains per population size estimated for DENV-2 was closer to that previously observed in rural regions in Thailand with less continuous endemic transmission (Fig. 4F).

All told, our phylogenetic analyses describe the emergence of a previously unrecorded genotype of highly divergent DENV-2 in at least ten different Cambodian provinces. This genotype greatly increased in prevalence following its first record in Cambodia in 2019 and caused infections in older-age individuals. The limited phylogenetic diversity observed from DENV-2 sequences suggest that the Cosmopolitan lineage (Genotype-II) arrived relatively recently to the region. Its high genetic divergence from previously reported genotypes in Cambodia may be associated with antigenic novelty that played a role in the 2019 epidemic.

### Mechanistic simulations of clade replacement with antigenic escape in an age-structured model recapture patterns witnessed in the data

Combined, our FOI analysis and genotyping suggest that the DENV-2 Cosmopolitan lineage (Genotype-II) caused a genotype replacement event in Cambodia, which likely played a part in the 2019 epidemic in the region. The higher burden of cases in older age individuals more likely to have some form of pre-existing dengue immunity additionally suggested that the Cosmopolitan genotype may be sufficiently antigenically divergent to overcome this immunity, resulting in symptomatic infection. To formally evaluate this hypothesis, we constructed a mechanistic, age-structured discrete time deterministic epidemic model in biweekly timesteps (*SI Appendix*) (58–60), with which we simulated diverse mechanisms hypothesized to underpin observed patterns in our data. We sought to explain both the general pattern of increasing average age of reported infection through time, as well as the high caseload and wide distribution in age range of cases associated with the 2019 epidemic (and, to a lesser extent, prior epidemic years). Previous work in Thailand shows similar patterns of drastically older age infections in more recent years, which the authors explain via a mechanism of increased detectability of tertiary or quaternary infections through time (23). In practice, this mechanism is similar to our extension of Ferguson et al. (1999)’s catalytic model for dengue, which incorporates waning multitypic immunity, as both approaches allow for case detection in individuals experiencing more than two dengue infections across their lifetime. We used our simulation model to evaluate whether these two mechanisms were functionally distinguishable and, if so, which best replicated Cambodian data.

For simplicity, we restricted ourselves to a three-serotype system and initiated simulations incorporating seasonal variation in transmission as estimated by TSIR (Fig. 1) and annual variation in FOI corresponding to national estimates (Fig. 3). As with our FOI analysis, we modeled secondary infections as reported cases for (H0) normal demographic simulations. We then compared these results against simulations of (H1) hypotheses that allowed for continuously increasing detectability of tertiary cases through time or (H2) hypotheses of genotype replacement paired with waning multitypic immunity and detectable reinfection within a serotype (Fig. 5). We also compared (H3) hypotheses of genotype invasion with no lineage-specific waning immunology or (H4) hypotheses of genotype invasion with continuous increasing tertiary case detection, again not specific to the invading lineage (Fig. S21). We focused our analyses on drivers of the 2019 epidemic but also conducted a subset of simulations to highlight insights gleaned from the 2007 epidemic in Cambodia (Fig. 5, *bottom*).

**Figure 5.**
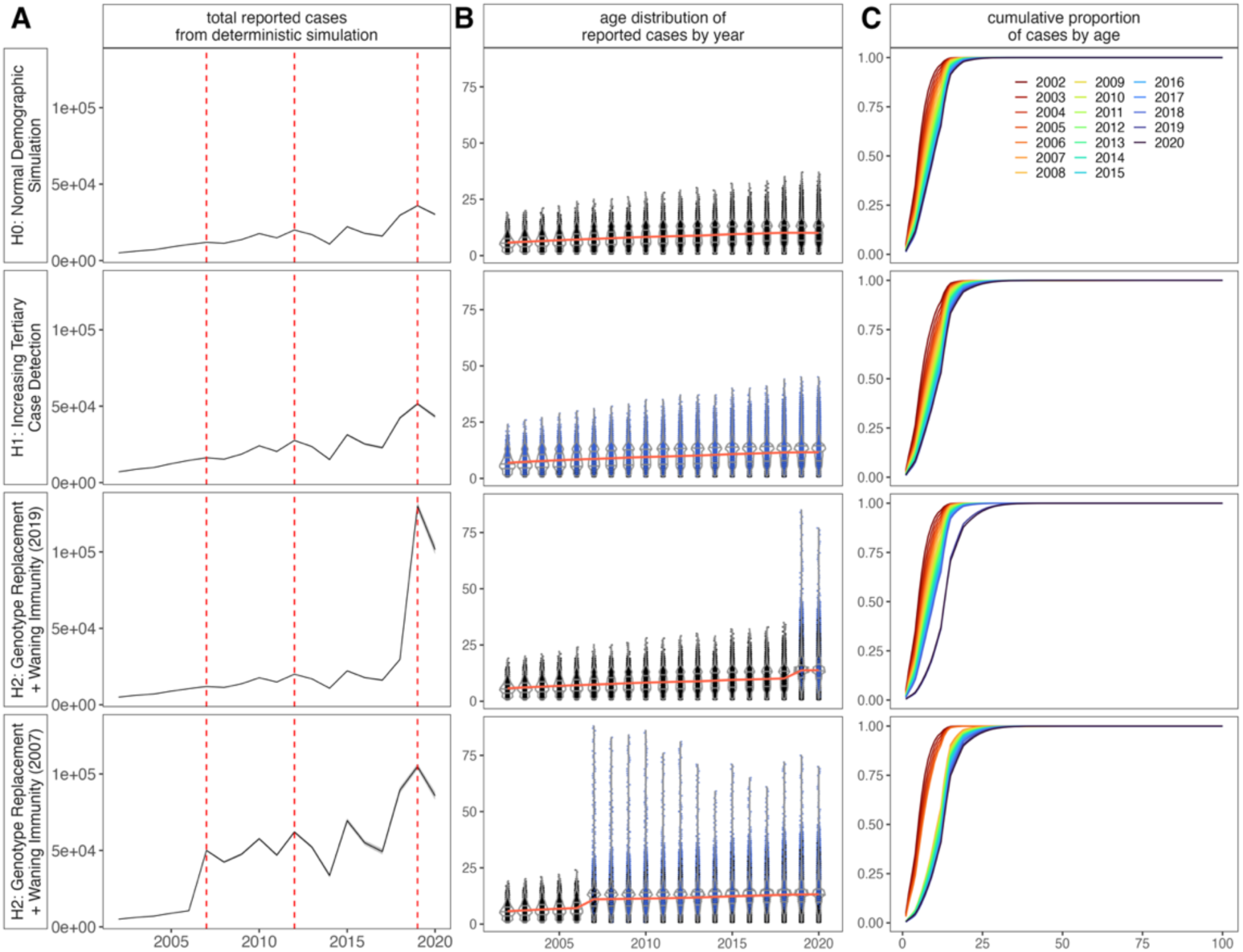
Simulations of genotype clade replacement recapitulate observed expansion in the age structure of reported dengue cases from Cambodian data. **A** Total reported cases (solid line = mean FOI; translucent shading = 95% confidence interval for FOI), **B** age distribution of reported cases by year (black = secondary; blue = repeat secondary or tertiary), and **C** cumulative proportion of reported cases by age, from deterministic model simulations indicated to the left of panel A.

All simulations successfully recovered the pattern of increasing mean age of reported infection, as witnessed in the data. This increase was steeper under (H1) assumptions of increasing detectability for tertiary infections (Fig. 5). Because we parameterized FOI from the national time series for Cambodia, for which the 2019 FOI was locally elevated (Fig. 3), all simulations also effectively reproduced a higher burden of cases in 2019. Critically, we found that sudden, sharp expansions in the age distribution of reported cases, as witnessed broadly across provinces in epidemic years for Cambodia (Fig. S15), were only possible when modeling some form of antigenic novelty for the invading lineage (H2; Fig. 5). Simulations of this novel antigenic invasion in 2007 (Fig. 5 H2, *bottom*) also demonstrated how the expanded age distribution should subsequently narrow in the years following clade replacement with immune evasion, again consistent with Cambodian data (Fig. S15). By contrast, (H1) hypotheses of constant increasing tertiary case detection through time will only continue to expand this distribution over longer time horizons. Importantly, modeling of genotype turnover had no demonstrable impact on the age distribution of cases in the absence of *specific* enhanced detectability for the invading clade, even when allowing for generally heightened tertiary case detection through time (Fig. S21). Thus, we conclude that only introduction of antigenically divergent genotypes (or, of course, serotypes) can be expected to generate this dramatic short-term expansion in the age distribution of cases.

Notably, our results were agnostic to classification of repeat infections with an invading lineage as tertiary detections (e.g. detection of DENV-2 infection following prior exposure to DENV-1 and DENV-4) vs. repeat detections within a serotype (e.g. detection of DENV-2 following prior exposure to a previous lineage of DENV-2). The epidemic consequences of this invasion were comparable, so long as enhanced detectability (e.g. antigenic escape) was specific to invading lineage.

## DISCUSSION

Drawing from two decades of national surveillance data and a more recent genomic cohort study, we queried the mechanisms that underly dengue virus transmission and drive periodic dengue epidemics in Cambodia. All told, our study highlights the complex interplay of climate, demography, immunology, and virology that dictates the dynamics of dengue virus disease in endemic settings.

Our investigations of climate effects on dengue transmission in Cambodia mirror those previously reported in Thailand (28, 29), Sri Lanka (50), and China (46), emphasizing the role of temperature, and—to a lesser extent—precipitation, in synchronizing annual dynamics in epidemic years. Consistent with prior work that highlights a role for El Niño in driving multiannual dengue cycles in other systems (29), we observed synchrony between reconstructed multiannual dengue cycles in Cambodia and the ONI; however, this synchrony appeared to peak during the robust 2015-2016 El Niño, which did not correspond to one of the three major dengue epidemic years in our dataset—though some signature of elevated amplitude for multiannual dengue cycles in northern Cambodia (adjacent to the Thai border) suggest a local epidemic may have occurred during this time. The muted impact of climate on dengue dynamics in Cambodia could reflect overall higher caseloads in association with elevated temperatures in more recent years (28), making any further transmission gains comparatively less dramatic. In addition, climate impacts on DENV transmission may be generally less discernable in Cambodia, which has reduced latitudinal variation (and corresponding climate differences) than other SEA countries, such as Thailand. Nonetheless, our analysis suggests that warmer temperatures play a role in driving epidemic dynamics for Cambodian dengue and likely contributed to high caseloads in 2019. As temperatures increase globally, dengue transmission is likely to accelerate, and variation in both timing of annual dengue transmission and multiannual peaks in caseload may become more homogenous.

In addition to climate, our data and corresponding analysis emphasize the importance of human demography and prior immunity in structuring dengue transmission globally. Consistent with studies conducted elsewhere (23–25), we observed a pattern of increasing mean age of reported dengue infection that we effectively recaptured by modeling a declining FOI through time—which we attributed to declining birth and death rates consistent with Cambodia’s demographic transition. Certainly, other mechanisms of demographic change (e.g. immigration and emigration) could also impact infection dynamics. Cambodia experienced a net negative migration rate across the time series investigated (61); as the majority of Cambodian migrants are young adults seeking employment in neighboring Thailand (62), it is possible that their emigration could lower the dengue FOI even further. Though the Cambodian migrant population comprises <0.5% of the total national population (61), migration rates can be 10-fold higher in border provinces and could have correspondingly elevated impacts in these regions (62).

The declining FOI through time is somewhat counterintuitive considering recent explosive dengue epidemics in Cambodia and across the globe. Nonetheless, we provide evidence for a high frequency of reported cases in older age individuals that underlies the high case burden in epidemic years. Our study is unique from previous investigations in recognition, not only of escalation of the mean age of reported dengue infection, but also of the expanding age range of reported disease. Indeed, symptomatic individuals >70 years in age during the 2019 epidemic are difficult to reconcile under assumptions by which reported cases correspond to clinically apparent secondary infections only. In our modeling framework, we take a novel approach to account for these infections by allowing for waning multitypic immunity and repeated symptomatic infections in older age individuals.

Recent analyses out of the Thailand system highlight a similar uptick in case reports among older individuals (23), which the authors attribute to increased detectability in tertiary and quaternary infections. The authors offer a series of hypothetical explanations for increasing case detectability through time: that tertiary and quaternary dengue infections might be more pathogenic (and therefore more detectable) due to an abundance of comorbidities in older individuals, that immunopathology may be exacerbated in older patients who experienced longer durations between repeat infections; or that waning multitypic immunity could allow for repeat infections in the oldest age cohorts (our hypothesis). Our analyses in the Cambodia system are not mutually exclusive with any of these explanations; however, we hypothesize that, were comorbidities or immunopathogenesis in older individuals driving patterns in the observed data, we would expect to see an hour-glass shape in symptomatic cases, with reduced reporting in middle-aged individuals who have progressed beyond secondary exposures but are at lower risk for both comorbidities and immunopathology. Instead, we see a gradual tapering in the age-frequency of infection, which expands in range with time (Fig. S15), consistent with the hypothesis of waning multitypic immunity. This latter hypothesis, when considered in association with genotype replacement, can also explain isolated expansion and subsequent contraction in the age range of reported cases witnessed in epidemic years, while the first two hypotheses cannot. In addition, hypotheses of heightened pathology in more frequent older age infections are difficult to reconcile in Cambodia with nationally-reported declines in the dengue mortality burden despite increasing caseload through time (15).

Our deterministic model simulations demonstrate how short-term spikes in the age distribution of reported cases, independent of the mean annual trend in the time series, can be achieved via introduction of an antigenically distinct lineage (either a divergent genotype within an established serotype, or a novel serotype) into an endemic, 3-serotype system. Though limited in scope, PCR-based sentinel surveillance data provide no support for serotype turnover associated with the 2019 dengue epidemic in Cambodia (15). By contrast, genome sequencing data from our own febrile cohort study offer support for a genotype replacement event. Recent analyses of the Thailand system link transitions in serotype dominance and clade replacement of genotype sublineages within a single serotype to epidemic magnitude (34, 35). In Thailand, the most severe epidemics were linked to clade replacement of a resident viral genotype by an evolutionarily fitter DENV lineage within the same serotype, resulting in a ‘selective sweep’, which subsequently reduced overall viral diversity and, consequently, diminished between-serotype antigenic differences (34). In the 2019 epidemic in Cambodia, emergence of the Cosmopolitan genotype, followed by serotypic and genotypic homogeneity in 2020-2022, are consistent with the dynamics of clade replacement within DENV-2. Indeed, prior clade replacements have been associated with epidemics in Cambodia (17, 18) and elsewhere (50, 54, 63–65).

As one limitation of our study, the bulk of our genotyping efforts were concentrated in peri-urban Kampong Speu province, allowing for the possibility that the clade replacement phenomenon documented here may be regionally localized, with consequentially limited impacts on national caseloads. Nonetheless, the predominance of the DENV-2 Genotype-V lineage in the mid-2000s has been well-documented across most provinces in Cambodia (66, 67), and our extended sequencing efforts in 2021-2022 suggest that the Cosmopolitan (Genotype-II) takeover covered a similarly widespread geographic extent. Additionally, reemergence of DENV-2 Cosmopolitan lineage has also been documented in neighboring Vietnam and associated with a 2019 epidemic (53), suggesting a more generalized regional phenomenon.

Our study faces additional limitations in that we were unable to obtain virus isolates and undertake subsequent antigenic cartography (68), which would be needed to resolve whether the emerging Cosmopolitan genotype (Genotype-II) in Cambodia is, indeed, antigenically distinct from previously circulating Genotype-V lineages of DENV-2 and, potentially, more antigenically similar to endemic heterotypic viruses. Indeed, because much of our inference is derived from serotype-agnostic national surveillance data, we are unable to rigorously evaluate the role of a possible genotype replacement event in particularly driving a high burden of disease in the oldest-age (>70 years) individuals in NDCP dataset. Nonetheless, the expansion of this lineage following the 2019 epidemic suggests some enhanced fitness for this genotype in our setting— though, whether mediated by increased transmissibility, immune escape, or both, is difficult to say. Our own febrile surveillance study demonstrates a significantly older mean age of infection for DENV-2 Cosmopolitan cases than for DENV-1 cases, which supports a hypothesis of invasion paired with immune escape. Critically, our simulation analyses demonstrate how the expanded age distribution of cases witnessed across epidemic years in Cambodia can only be recovered under assumptions of invasion or expansion with lineage-specific waning immunity.

Several published studies offer regional explanations for the global dengue phenomenon of 2019. Recent analyses from Brazil, for example, argue that a low FOI in 2017 and 2018 resulting from new public health interventions and behavioral modifications implemented in the wake of the Zika virus epidemic drove a resurgence in cases in 2019 and an expansion of specific lineages of DENV-1 and DENV-2 that had been circulating cryptically for much of the past decade (69). Independent work in the same region indicates that the DENV-2 lineage responsible for the Brazilian outbreak additionally caused a clade replacement event (64).

Dengue surged worldwide in 2019, though the factors driving this surge appear to be somewhat heterogenous across ecosystems. Nonetheless, our study lends support for the role of climate in synchronizing epidemic dynamics across landscapes; it is possible that optimal climate conditions may facilitate expansion of low prevalence lineages with intrinsic fitness advantages (e.g. higher replication rate, shorter incubation period (70)) over resident genotypes, thus driving epidemics. Previous modeling studies have suggested that invading dengue lineages may circulate at low prevalence in a host population for many years prior to detection, expansion, and displacement of resident clades in the same serotype (71).

Here, we propose the emergence of the DENV-2 Cosmopolitan genotype (Genotype-II), coupled with a subtle climate-driven increase in FOI, and overlaid on the background of an aging population with waning multitypic immunity, as one possible explanation for Cambodia’s largest recorded dengue epidemic to date. Our study emphasizes the extraordinary dearth of publicly available DENV sequence data for SEA; indeed, DENV is sequenced so infrequently in Cambodia that it is impossible to know whether 2019 truly marked the first year of DENV-2 Cosmopolitan introduction to the country, or simply the year of intensified expansion and consequential epidemic dynamics. More broadly, our work illustrates the importance of the combined forces of climate, demography, immunology, and virology in driving increasingly severe dengue epidemics. As the global burden of dengue continues to expand, ongoing serological and genomic surveillance is needed to improve epidemic forecasting in Southeast Asia and around the world.

## METHODS

### Data

Detailed descriptions of NDCP case data, climate data, and Kampong Speu febrile surveillance data used in these analyses are reported in the *SI Appendix*.

### Climate analyses

We used GAMs to quantify intra- and interannual trends and identify any years that might be considered climatic anomalies, defined as a statistically significant partial effect of a single year on the long-term time series for temperature and precipitation (Fig. S1-4; Table S1; *SI Appendix*). In addition, we reduced temperature and precipitation time series by province into z-scores for visualization (Fig. S5-6).

### Climate-informed TSIR modeling

Using the R package tSIR (43), we fit time series Susceptible-Infected-Recovered models (43–45) to the province level dengue data for the three inter-epidemic periods (2002-2006, 2008-2011, and 2013-2018), compiled at biweekly intervals corresponding to the generation time of the pathogen (*SI Appendix*). Using TSIR transmission parameters, we projected cases in subsequent epidemic years (2007, 2012, 2019) by province. We then followed Wagner et al. 2020 (50) to estimate the factorial increase in reconstructed susceptible population needed to recover case counts for the three epidemic years.

We next used cross correlation analysis to determine the optimal lag between biweekly mean temperature and total precipitation versus transmission at the province level; we estimated a median 3.5 month lag for temperature and 1 month lag for precipitation, consistent with prior analyses (Table S3; *SI Appendix*) (50). We then constructed a suite of regression models (including GAMs) for each interepidemic period, incorporating a response variable of the log of biweekly transmission with the corresponding predictors of optimally lagged biweekly mean temperature and total precipitation per province (46, 50) (Fig. S8-10; Table S4). We used lagged temperature and precipitation from each epidemic year in each province in a fitted GAM to project climate-informed transmission rates for 2007, 2012, and 2019 (*SI Appendix*). We then simulated TSIR using climate-informed transmission rates to recover epidemic year predictions of province-level caseload. Because climate-derived transmission rates still failed to recover epidemic peaks, we allowed another susceptible amplification term to determine the relative increase in susceptible population needed to recover epidemic dynamics even after accounting for climate (Table S5).

### Wavelet analyses

We next used wavelet decomposition in the R package ‘WaveletComp’ (51) to explore annual and multiannual periodicity in dengue epidemics and climate variables. We converted biweekly case totals by province from the NDCP data into incidence rates per 100,000 population, then used a Morlet wavelet with nondimensional frequency (𝜔 = 6) to extract, detrend, and reconstruct annual cycles in dengue incidence with a maximum period of two years and multiannual cycles with periods ranging from two to 20 years (Fig. 2AB) (28, 29). We additionally calculated the average wavelet power (squared amplitude, here averaged across all significant wavelets within a specified timestep) for significant cycles within each biweekly timestep for annual and multiannual cycles per province (Fig. S11AB).

We investigated synchronicity in dengue incidence across space by computing the Pearson’s correlation coefficient (𝜌) and the mean cross-wavelet power spectrum between province pairs, using the annual raw incidence and reconstructed annual and multiannual cycles (the latter averaged over a sliding 5-year interval) (Fig. 2CD, S11CD; *SI Appendix*). We constructed a GAM to identify statistical correlates of high synchronicity pairs (Fig. S12; Table S6; *SI Appendix*). To investigate synchrony between dengue and climate, we computed mean cross-wavelet power between biweekly dengue incidence and mean temperature (Fig. S13A) and total precipitation (Fig. S13B) per province, then repeated between reconstructed multiannual dengue cycles over a 5-year interval and the same two climate variables, again per province (Fig. S13CD). Finally, we reconstructed *monthly* multiannual dengue cycles per province to compute average cross-wavelet power with the Oceanic Niño Index (ONI), a time series quantifying the intensity of the El Niño Southern Oscillation (Fig. S13E). Lastly, we extracted the mean period from reconstructed multiannual dengue cycles by province and nationally, to compare with demographic data (birth rates, death rates, population size (61)) (Fig. S14).

### Increasing age of reported infection and FOI estimation

To quantify interannual trends in the mean age of reported DENV infection, we fit a GAM with a response variable of age to a fixed predictor of the interaction of year and province, with a random effect of province, allowing both slope and y-intercept to vary by locality. We visualized trends across the age distribution of reported cases by province (Fig. S15) and summarized nationally (Fig. 3AB; Table S7).

To estimate annual FOI for DENV in Cambodia from 2002-2020, we applied the model developed by Ferguson et al. 1999 (36) and Cummings et al. 2009 (24) to age-structured incidence recovered from the NDCP data, at the province level, assuming reported cases to represent secondary infections and individuals in the dataset to experience exposure to multiple DENV serotypes across their lifetimes. We allowed for a unique FOI across each year in the time series but first assumed constant FOI across all age cohorts within a year. We estimated one FOI per year per province (excepting Tboung Khmum), in addition to a summary national FOI, for all years in the dataset and extending back to the year of birth for the oldest individual in the first year of the corresponding data subset (40 years for national data) (Fig. S16; Table S8; *SI Appendix*). We estimated mean FOI per serotype assuming four circulating dengue serotypes and plotted FOI estimates in comparison to national reported birth and death rates (Fig. 3C) (61).

After fixing FOI by province, we followed prior work (24) to estimate FOI modifiers by age, shared across all provinces. We fit nineteen age-specific modifiers to the fixed FOI values, corresponding to eight age classes in first decade of the dataset and eleven in the second decade, where the age distribution was larger (Table S8). We then modified our model to allow waning from multitypic back to monotypic immunity (𝜎), such that older individuals could experience renewed symptomatic infections (*SI Appendix*). As with age-modification terms, we estimated waning multitypic immunity shared across all provinces but allowed 𝜎 to vary annually. We compared fits of FOI-only, FOI with age modification, FOI with waning multitypic immunity, and FOI with both age modification and waning multitypic immunity models to the data (Table S8), then simulated the resulting accumulation of cases with age from the best fit model at national (Fig. 3E) and province levels (Fig. S17).

### Viral sequencing

mNGS was applied to RNA extracted from 239 serum samples collected from patients reporting with symptoms in our 2019-2022 febrile cohort study in Kampong Speu province, with an additional subset of 11 samples sourced from five nearby provinces (40). A further 20 samples were received for RNA extraction and mNGS from six provinces (including 8 from Kampong Speu) in part with 2021-2022 NDCP surveillance (Table S9; Fig. 4AB). See *SI Appendix* for detailed methods for library preparation, sequencing, and consensus genome generation. We successfully generated 192 full or near-full genome sequences from the original 270 samples, which we submitted to NCBI (63 DENV-1, 120 DENV-2, and 9 DENV-4). All contributed genomes were >10,000 bps in length and had a maximum of 90 Ns (corresponding to <1% of the DENV genome). Samples for which full genome recovery was not possible were identified to genotype using BLAST (72) and visualized in Fig. 4AB (Table S9).

### Phylodynamic and phylogenetic analysis

See *SI Appendix* for details of sequence selection for Bayesian phylodynamic timetrees and maximum likelihood phylogenies. After selection, sequences were aligned by serotype in the program MAFFT (73), and the best fit nucleotide substitution model for each serotype was evaluated in ModelTest-NG (74). Using the best-fit model (GTR+I+G4 in all cases), we built a Bayesian phylogenetic tree for each serotype in BEAST 2 (57), incorporating the date of sample collection for each sequence (or the midpoint of the collection year if date was not reported), and specifying a strict molecular clock rate of 7.9x10^-4^ s/s/y (8) and a Coalescent Bayesian skyline prior. We ran Markov Chain Monte Carlo chains for 150 million iterations, logging results every 10,000 iterations. After chains completed, we removed the initial 10% of iterations as burn-in and evaluated parameter convergence (ESS≥ 200) in Tracer v1.6. We summarized resulting phylogenetic trees in TreeAnnotator and visualized summary trees in ggtree (75) (Fig. 4CD; Fig. S18, S19). Serotype-specific maximum likelihood phylogenetic trees were constructed in RAxML (76). Twenty ML inferences were made on each original alignment and bootstrap replicate trees were inferred using Felsenstein’s method (77), with the MRE-based bootstopping test applied after every 50 replicates (78). Resulting phylogenies were visualized in ggtree (75) (Fig. S18).

We followed Salje et al. 2017 (8) to calculate the proportion of sequences within each DENV serotype that could be attributed to the same transmission chain on our Bayesian timetrees, defined as having an MRCA within the past six months in the same season (see Fig. S20 for alternative MRCA cutoffs). We compared the proportion of DENV-1 vs. DENV-2 sequences sharing a transmission chain to the Euclidean distance separating the GPS coordinates for each sequence pair (Fig. 4E). We then computed the total effective number of transmission chains (the reciprocal of the proportion of sequences sharing a chain) in our 2019-2022 Kampong Speu sequence dataset for DENV-1 and DENV-2 at the World Bank reported population density for Kampong Speu province (61) (Fig. 4F).

### Mechanistic modeling of age-structured dengue dynamics

Finally, we constructed a mechanistic, age-structured discrete time deterministic model in biweekly timesteps (58–60) to simulate three-serotype dengue dynamics in a population demographically structured to mimic that of Cambodia over the past half-century (*SI Appendix*). We compared simulations under (H0) baseline demographic assumptions with simulations of (H1) increasing tertiary case detections through time and (H2) genotype invasion and clade-replacement with waning immunity, timed both in 2019 and in 2007. We additionally simulated novel genotype invasions under (H3) assumptions of no antigenic escape, or (H4) lineage-agnostic increasing tertiary case detection (Fig. S21). Finally, we fit the Ferguson-Cummings four-serotype catalytic model to the simulated data for each hypothesis to recapture the input FOI (Fig. S22). Holding FOI constant, we estimated a time-varying signature of waning monotypic immunity for all simulated time series (Fig. S22).

## Data Availability

All genome sequence data from this study have been submitted to the NCBI Sequence Read Archive under Bioproject ID PRJNA681566. Consensus DENV sequences are also available in GenBank, under the following accession numbers: DENV-1: OK159935-OK159976, OL411495-OL411499, and OL412140, OL412678, and OL412703; DENV-2: OL414717-OL414765, OL412740, OL420733 and OL435143; DENV-4: MZ976858-MZ976860).

https://github.com/chanzuckerberg/idseq-workflows

https://artic.readthedocs.io/en/latest/

https://docs.google.com/document/d/1RtNQc1D4or_ys7OxCCBjh4SDIdy7JaI4IE7if8EkHgE/edit

https://github.com/brooklabteam/cambodia-dengue-national

## ACKNOWLEDGEMENTS

This research is supported by the Division of Intramural Research at the National Institute of Allergy and Infectious Diseases at the National Institutes of Health and the Bill and Melinda Gates Foundation [OPP1211806, OPP1211841]. We thank patients and families of Kampong Speu District Referral Hospital who participated in this study, members of the National Dengue Control Program not listed in the author byline, and the Provincial Health Department of Kampong Speu province in Cambodia. We thank Brian Moyer and the NIAID Office of Cyberinfrastructure and Computational Biology (OCICB) for assistance in improving the cyberinfrastructure of our Cambodian field sites. We thank employees at the Chan Zuckerberg Biohub and Chan Zuckerberg Initiative not listed in the author byline. This work was completed with resources provided by the University of Chicago Research Computing Center.

