## Supplementary Appendix for "Climate, demography, immunology, and virology combine to drive two decades of dengue virus dynamics in Cambodia"

**This PDF file includes:**

Supporting text
Figures S1 to S22
Tables S1 to S10
SI References

### Supporting Information Text

### Supporting Text 1: Data Sources

#### *1.1 National Dengue Control Program data*

We obtained a 2002-2020 time series of age-structured dengue cases reported at the national level from clinicosyndromic surveillance efforts administered by the National Dengue Control Program (NDCP) of the Cambodian Ministry of Health. Since inception of the national surveillance system in 2002, each Cambodian province reports cases on a monthly basis to the national authorities who compile these reports for public health use and reporting to the World Health Organization. The date of each case and the corresponding age and gender of each patient are reported, in addition to the province in which the case was diagnosed (25 total). We binned cases to every-two-week (biweekly) intervals for application to TSIR modeling and summarized by age class within each year for application to FOI models. Tboung Khmum was created only recently (2016) and, correspondingly, eliminated from both TSIR and FOI analyses.

#### *1.2 Spatially-resolved climate data*

Climate variables, including daily mean temperature ($^0$C) and total daily precipitation (mm) from 2002 to 2019 were downloaded from the ERA5 Daily Aggregates dataset for each Cambodian province (N=25) using Google Earth Engine (1). Data were not available for 2020. The spatial resolution for the ERA5 data set is approximately 31 km. We aggregated climate data to the size of each province using the Cambodia Administrative Boundaries Level 1 shapefile (2) during the extraction process, then computed the mean temperature ($^0$C) and total precipitation (mm) over biweekly intervals, from January 1, 2002 to December 31, 2019, while accounting for leap years (Fig. S1-S2).

#### *1.3 Kampong Speu febrile cohort study*

This study was approved by the National Ethics Committee on Human Research and the National Institutes of Health (NIH) Institutional Research Board. Written informed consent was obtained from the individual participant or the parent or guardian of the child participants enrolled in this study. This study was registered at clinicaltrials.gov as NCT04034264 and NCT03534245.

Nursing staff at the Kampong Speu (KPS) Referral Hospital identified, consented, enrolled, and collected demographic data from study participants. Participants, aged 6 months to 65 years of age, presented to the outpatient department with a documented fever of 38°C or greater in the previous 24 hours. Participants with clinical symptoms and signs consistent with dengue were first screened for infection using SD Bioline DengueDuo rapid tests for NS1 antigen, pan-dengue IgM and IgG. Sera was collected and processed as described elsewhere for RNA extraction, and confirmatory qRT-PCR testing for DENV-1 – 4 was performed for rapid-test positive participants (3).

**Supporting Text 2: Climate analyses**

*2.1 Generalized additive models (GAMs)*
We explored the extent to which epidemic years could be considered climatic anomalies using four discrete generalized additive models (GAMs) in the mgcv package in R (4).

The first GAMs incorporated a response variable of biweekly mean temperature or total precipitation, at the province level, with a fixed predictor of the interaction of year with province, a cyclic smoothing spline by biweek of year, and a random effect of province. Each GAM fit 25 distinct slopes and 25 distinct y-intercepts to describe the interannual trend in temperature or precipitation for each province, while controlling for intra-annual variation (Fig. S3-S4; Table S1).

We next constructed two GAMs with the same response variables but incorporating predictor variables of year as a random effect and biweek as a cyclic smoothing spline to identify specific years that significantly deviated from mean climate trends, representing climatic anomalies. We computed 95% confidence intervals by standard error on the partial effect of each year on the overall trend while holding all other variables constant. Years for which 95% CIs of the effect size did not overlap zero were considered significant climate anomalies, with anomalously warm years corresponding to those with significantly positive effect sizes and anomalously cool years corresponding to those with significantly negative effect sizes (Fig. S5-S6; Table S1).

**Supporting Text 3: The TSIR Model**

*3.1. General theory behind the TSIR model*
One of the simplest and most celebrated epidemic models, the Susceptible-Infectious-Recovered (SIR) model describes the dynamics of infectious disease transmission through a well-mixed host population, driven by rates of transmission ($\beta$) and recovery ($\gamma$) from infection, coupled with host demographic processes of birth ($B$) and death $\mu$ (5, 6). In its most standard form, the SIR model assumes that all individuals are born susceptible and that immunity after infection is lifelong:

$$\frac{dS}{dt} = B + \frac{\beta SI}{N} - \mu S$$
$$\frac{dI}{dt} = \frac{\beta SI}{N} - \gamma I - \mu I$$
$$\frac{dR}{dt} = \gamma I - \mu R$$

[1]

In standard annotation, the transmission rate ($\beta$) and the proportion of the population infected ($\frac{I}{N}$) can be characterized simply as the 'force of infection' ($\lambda$), the rate at which susceptibles become infected. One widely-used subclass of SIR models, the time series Susceptible-Infected-Recovered model, or the TSIR, was developed to simplify the process of parameter estimation in the fitting of SIR models to time series data, particularly for perfectly-immunizing childhood infections (e.g. measles) (7–10). The TSIR depends on two key assumptions: (*i*) that the pathogen infectious period is equal to the data sampling interval (classically, biweekly for measles) and (*ii*) that, over lengthy, 10-20 year time horizons, the sum of infected cases should roughly equal the sum of births for highly infectious childhood diseases for which all individuals are expected to eventually be exposed. The model assumes no overlapping generations of infection, and, following this logic, approximates the number of infections in a given timestep as the product of the susceptible population multiplied by the force of infection in the preceding timestep (equation [2]), where the homogeneity parameter ($\alpha$) captures epidemic saturation and serves as a correction factor in the process of switching the model from continuous to discrete time:

$$S_{t+1} = B_t - S_t - I_t$$
$$I_{t+1} = \beta_{t+1} S_t I_t^{\alpha}$$

[2]

Given a time series of infected cases of childhood disease, the TSIR framework can thus be implemented by first reconstructing the susceptible population. In this process, a regression model is fitted between cumulative cases and cumulative births. If assumed to be equal, the slope of this regression model gives the reporting rate through time ($\rho_t$), and the residuals from this slope, ($Z_t$), represent heterogeneity in the susceptible population beyond the average. The mean of the susceptible population across the time series, ($\bar{S}$), can then be inferred from profile likelihood, and the time-varying regression rate ($\beta_t$), along with the homogeneity parameter ($\alpha$) can be estimated using a generalized linear model after the following form:

$$\log I_{t+1} = \log \beta_{t+1} + \log(Z_t + \bar{S}) + \alpha \log I_t$$

[3]

A complete description of the TSIR model and underlying algorithms can be viewed in Finkenstädt and Grenfell 2000 (9). For the purposes of our analysis, we implemented the TSIR model using the R-package, tSIR, from Becker and Grenfell 2017 (7). Fixed and estimated parameter values for discrete fits to three subsets of dengue time series (2002-2006, 2008-2011, and 2013-2018) are available for viewing in Supplementary Table S2.

*3.2 Details of province level TSIR for Cambodia dengue*

To fit province level TSIR models to each inter-epidemic period, we divided nationally reported birth rates for Cambodia (11) evenly among biweeks of each year and scaled them spatially by the relative population size of each province. For each inter-epidemic period, we reconstructed the susceptible population per province from the regression of cumulative cases on cumulative births, using either a Gaussian or a linear regression, depending on which provided the better fit to each data subset (Table S2). Provinces Ratanak Kiri and Mondul Kiri were excluded from analysis due to poor performance of regression models for susceptible reconstruction (Table S2), and province Tboung Khmum was excluded because the time series only initiated in 2016. For all provinces retained for TSIR analysis, we estimated 26 biweekly intra-annual transmission rates ($\beta$) and one homogeneity parameter ($\alpha$) for each inter-epidemic period from the data.

To implement a climate-informed TSIR model, we expressed the log of the biweekly transmission rate as a function of the optimally lagged biweekly mean temperature and total precipitation for the province and year in question. In keeping with prior work for dengue (12), we conducted cross correlation analysis to determine the optimal lag between the time series of biweekly mean temperature and total precipitation on transmission at the province level. We considered only lag times of up to one year in which the climate predictor led the transmission rate and allowed the optimal lag to vary by province and inter-epidemic period. We found that the optimal time lag between peak climate predictor and peak dengue transmission ranged from 0.5 to 5 months (1 – 11 biweeks) for temperature (median = 3.5 months) and from 0.5 to 9 months (1 – 18 biweeks) for precipitation (median = 1 month), consistent with prior analyses (12) (Table S3).

Using these optimal lags, we next constructed a suite of regression models for each interepidemic period, incorporating a response variable of the log of biweekly province level transmission with the corresponding predictors of optimally lagged biweekly mean temperature and total precipitation for the province in question (12, 13) (Fig. S8-10; Table S4).

We investigated three forms of regression analyses by which to best represent the time-varying transmission rate, exploring a standard linear mixed effects regression; a linear mixed effects regression for the precipitation term, coupled with a Brière function for temperature; and a generalized addition model (GAM) (4), which we ultimately selected for the results reported in the main text.

The linear mixed effect regression (output visualized in Fig. S8A, S9A, S10A), took the following form:

$$\log \beta_{t,i} = \alpha_0 + B_1 T_{lag_{t,i}} + B_2 P_{lag_{t,i}} + \mu_{0,i} + \varepsilon_{t,i} \quad [4]$$

where $B_1$and $B_2$ represent the slopes of the fixed predictors (respectively, lagged temperature $T_{lag}$ and lagged precipitation $P_{lag}$) for a specific province $i$ at time $t$, $\mu_{0,i}$ is the province-specific random intercept, and $\varepsilon_{t,i}$ is a normally distributed error term.

Following (12), we also explored a combined linear mixed effects regression for lagged precipitation and a Brière function for lagged temperature, such that the temperature term in equation [4] was replaced with the following function:

$$f_{Brière} T_{lag_{t,i}} = c(T_{lag_{t,i}} - T_{min})(T_{max} - T_{lag_{t,i}})^{1/2} \quad [5]$$

Mirroring previous work (14), we embraced maximum flexibility in delineating the relationship between climate predictors and biweekly transmission using a GAM in which the time-varying climatological covariates were fit as smooth splines (Fig. S8C, S9C, S10C:

$$\log \beta_{t,i} = s\left[T_{lag_{t,i}}\right] + s\left[P_{lag_{t,i}}\right] + \mu_{0,i} + \varepsilon_{t,i} \quad [6]$$

where the $s$ terms represent smoothing splines, $\mu_{0,i}$ is the province-specific random intercept, and $\varepsilon_{t,i}$ is a normally distributed error term. Because all models were equivalently significant for all three inter-epidemic periods (Supplementary Table S4), we selected the GAM to allow for maximal flexibility in capturing subtle variation across provinces and timesteps while projecting epidemic year transmission rates for TSIR prediction.

### Supporting Text 4: Wavelet Analyses

#### *4.1 Detailed methods for evaluation of synchronicity in dengue incidence between provinces*

We investigated synchronicity in dengue incidence across space by computing the Pearson's correlation coefficient ($\rho$), as well as the mean cross-wavelet power spectrum between province pairs, using the annual raw incidence and the reconstructed annual and multiannual cycles (Fig. 2CD, S11CD). All results for annual cycles were qualitatively similar to those for annual raw incidence and are, therefore, not reported here (Fig. 2C).

For annual incidence, we computed $\rho$ for each province pair combination in yearly timesteps, then calculated the annual average $\rho$ for each focal province compared against all other provinces (Fig. 2C). For multiannual cycles, we computed $\rho$ for all pairwise province combinations across a sliding 5-year window. As with annual incidence, we then calculated the average $\rho$ for each focal province per year, compared against all other provinces. Because 5-year cycles overlapped, $\rho$ was averaged over multiple overlapping comparisons for each pairwise combination for all but the first and last year in the time series (Fig. 2D).

For another measure of synchronicity, we additionally computed the cross-wavelet power spectrum for all province pairs, using both annual incidence rates in yearly timesteps and multiannual reconstructed cycles in 5-year intervals. As with the Pearson's correlation coefficient, we averaged the output of these analyses (here, significant values for cross-wavelet power) for each focal province, as compared with all other provinces, at the appropriate timestep (Fig. S11CD).

To identify statistical correlates of high synchronicity years and localities, we constructed a GAM with a response variable of $\rho$ from the annual incidence comparison (Fig. 2C); a fixed predictor of the interaction of focal province and geographic distance to the province under comparison; smoothing predictors of biweekly mean temperature, total precipitation, and mean population size for the focal province; and a random effect of year (Fig. S12; Table S6).

### Supporting Text 5: Age distribution of cases and the FOI age-cumulative incidence model

Methods for estimating the force of infection (FOI, $\lambda$) from age-structured serological data for perfectly immunizing infections have been long-established and are well-described (15–19): these models demonstrate that the proportion of individuals seropositive in a given age class can be approximated by age-specific FOIs that accumulate across the duration of time spent within the corresponding age class—akin to a survival model under variable age-specific hazards of mortality. Estimation of age-structured seroprevalence is more complex for dengue, as a result of the dynamics of four co-circulating serotypes, for which infection results in longterm homotypic immunity but enhanced susceptibility to heterotypic serotypes. More recent work additionally suggests that secondary infections with homotypic serotypes may be possible following primary infections, provided sufficient phylogenetic distance between the genotypes responsible for primary and secondary infections (20).

*5.1. Multi-typic exposures with life-long immunity*

For lifelong immunizing childhood infections for which all individuals are expected to experience infection at some point in their lifetime, the hazard of exposure will compile cumulatively with increasing time since birth (e.g. with age), making time and age interchangeable units. As a result, data describing the age-distribution of exposures can be used to estimate the force of infection (as it varies with time or age or both) in a given system.

Ferguson et al. 1999 (21) present a system of equations (PDEs) describing the dynamics of a multitypic dengue infection with rates in terms of time, $t$, and age, $a$. Ferguson et al.1999 then derive equivalent expressions describing the time-and-age-dependent population of susceptibles ($x$), the time-and-age-dependent population of individuals exposed to only a primary infection with serotype $i$ ($z_i$), and the time-and-age-dependent population of individuals experiencing any multitypic (2+ exposures) infection ($z_m$):

$$x(a,t) = e^{-\int_0^a \sum_i \lambda_i(a-\tau,t-\tau)d\tau} \tag{7}$$

$$z_i(a,t) = \left(e^{-\int_0^a \sum_{k\neq i} \lambda_k(a-\tau,t-\tau)d\tau}\right)\left(1 - e^{-\int_0^\infty \lambda_i(a-\tau,t-\tau)d\tau}\right) \tag{8}$$

$$z_m(a,t) = 1 - x(a,t) - \sum_i z_i(a,t) \tag{9}$$

In equation (7) – (9), the term $\tau$ reflects the inherent confounding between time $t$ and age $a$. The two variables change at the same rate (i.e. $\frac{dt}{da} = 1$) and therefore once an individual is born, the difference between their age and the current "time" remains fixed and can be tracked with a single time dependent variable.

Equation [8] describes the population of individuals exposed to only a primary infection with serotype $i$ ($z_i$) and can be read as the product of two probabilities:
(the probability of avoiding infection with all serotypes except for $i,$ up to time $t$).

× (the probability of not avoiding infection with serotype $i$)

Using equation [7], this expression can also be rewritten as:

$$z_i(a,t) = x(a,t)\left[e^{\int_0^a \lambda_i(a-\tau,t-\tau)d\tau} - 1\right]$$

[10]

Following Cummings et al. 2009 (22), we first estimate a time-varying, annual FOI for our Cambodian dengue system, then later add in variation by age class shared across all years and provinces in the dataset.

Cummings et al. 2009 (22) discretized the Ferguson system shown above, creating a piece-wise solution whereby they estimate an annual mean FOI ($\bar{\lambda}$) representative for all serotypes (because the available data are not serotype-specific, serotype-specific FOIs, $\lambda_i$, cannot be distinguished). Following Cummings et al. 2009 (22), the integrand in equation [7] can be reformulated as:

$$\int_0^a \sum_i \lambda_i(a-\tau,t-\tau)d\tau = \sum_0^a N\bar{\lambda}(a-\tau,t-\tau)\Delta\tau$$

[11]

where $N$ corresponds to the number of circulating dengue serotypes in the system and $\Delta\tau$ corresponds to the duration of time acted on by each $\bar{\lambda}(a-\tau,t-\tau)$, here, for simplicity, always held constant at one year.

Following on above, the second integrand in equation 8 can also be reformulated as:

$$\int_0^a \lambda_i(a-\tau,t-\tau)d\tau = \sum_0^a \bar{\lambda}(a-\tau,t-\tau)\Delta\tau$$

[12]

where, again, $\Delta\tau$ corresponds to the duration of time acted on by each $\bar{\lambda}(a-\tau,t-\tau)$, here held at one year.

We first followed Cummings et al. 2009 (22) to fit the above model to our dataset, estimating 40 distinct values for $\bar{\lambda}(t-\tau)$, one for each year from 1981-2020, beginning in the birth year (1981) of the oldest individual (22 years) in the first year (2002) of the dataset and extending through the last year of data for each province. We assumed 4 endemic circulating serotypes (e.g. N=4) in the system, as has been previously reported for Cambodia (23). Again, following Cummings et al. 2009 (22), we subsequently estimated 40 $\bar{\lambda}(t-\tau)$ paired with 19 age-specific variations on the annual $\bar{\lambda}(t-\tau)$ , 8 of which were shared across all provinces and years up to 2010 and 11 of which were shared across all provinces and years after 2010 where the age distribution of cases was larger. .

*5.2. Multitypic exposures with waning immunity*

Because we observed a sharp increase in the number of dengue cases reported in older (50+ years) individuals in the later years of our dataset, we next extended the model presented in Ferguson et al. 1999 (21) to include a rate of waning multitypic immunity, which allowed for re-infection with the same serotype ($i$) in later age classes.

We can conceptualize our new system in the following box model:

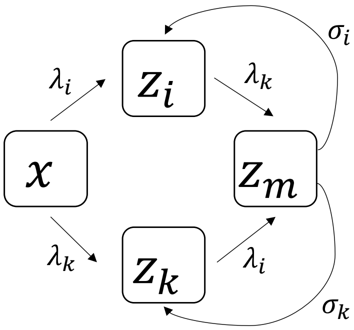


The above diagram assumes two circulating serotypes (represented with subscripts $i$ and $k$ , keeping with Ferguson's notation). Additional states could be added if additional serotypes were at play. Then we would use Ferguson's exact notation where $i$ refers to the focal strain and $k \neq i$ is an index representing all of the other strains. Here, $\sigma_i$ and $\sigma_k$ represent waning from a multitypic exposure state back to a homotypic exposure state ($z_i$ or $z_k$), allowing for re-exposure to $z_m$ and presentation as a reported case. For simplicity, we assume these rates to be constant across age and time. With the exception of the $\sigma$ terms, this model is identical to that presented in Ferguson et al. 1999 (21).

We express the first two terms in our system of differential equations as:

$$\frac{dx(a,t)}{dt} = -x \sum_i \lambda_i(a, t-a)$$

[13]

$$\frac{dz_i(a,t)}{dt} = x \sum_i \lambda_i(a, t-a) - \sum_{k \neq i} \lambda_k(a,t) + \ \sigma_i z_m(a,t)$$

[14]

where $z_i(a,t)$ represents the proportion of individuals that demonstrate history of homotypic infection with single strain $i$.

From [13], we can then solve directly for $x(a,t)$:

$$x(a,t) = Ce^{-\int_0^a \sum_j \lambda_j(a-\tau, t-\tau)d\tau}$$

[15]

where $j \in \{i,k\}$. We can then solve for $C$ (the integration constant) under the assumption that the entire population is born susceptible, $x(0) = 1$. From this, we determine that $C = 1$, revealing that the susceptible population is represented by the same expression previously shown for the system without waning immunity in equation 7 above:

$$x(a,t) = e^{-\int_0^a \sum_j \lambda_j(a-\tau, t-\tau)d\tau}$$

[16]

Following Cummings et al. 2009 (22) and using Cambodia data which lack serotype-specific specifications, we can estimate the mean FOI per serotype, assuming $N$ circulating serotypes in our system (we model this system under assumptions of N=4):

$$x(a,t) = e^{-\int_0^a N\bar{\lambda}(a-\tau, t-\tau)d\tau}$$

[17]

Following Ferguson, we can now derive an expression for $z_i(a,t)$. This expression should sum the probabilities of the three disparate routes by which an individual can enter the primary exposure state ($z_i$), as highlighted in the diagram below—either progressing directly from $x$ to $z_i$ (**red**) or achieving $z_m$ and then waning back into $z_i$ (**green** or **blue**):

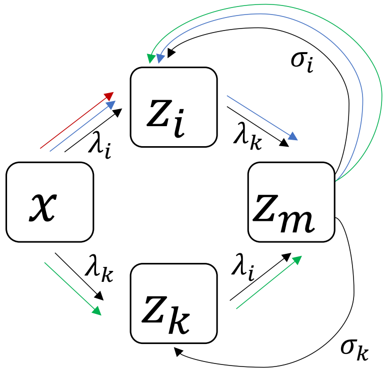


We write this new expression as the summed probabilities of the three pathways, with the second and third pathways combined to describe the product of the sequential probabilities of achieving $z_m$ by any means and then waning back into $z_i$:

$$z_i(a,t) = \left(e^{-\int_0^a \sum_{k\neq i} \lambda_k(a-\tau,t-\tau)d\tau}\right)\left(1 - e^{-\int_0^\infty \lambda_i(a-\tau,t-\tau)d\tau}\right) + \left(1 - e^{-\int_0^a \lambda_i(a-\tau,t-\tau)+\lambda_k(a-\tau,t-\tau)d\tau}\right)\left(e^{-\int_0^a \sum_{k\neq i}\sigma_k d\tau}\right)\left(1 - e^{-\int_0^a \sigma_i d\tau}\right)$$

[18]

The summation term included with $\sigma_k$ allows for the possibility of including greater than two serotypes by which an individual could wane out of the multitypic exposure state.

After Cummings et al. 2009 (22), we can again discretize the system and estimate the average rate of waning immunity across all serotypes, $\bar{\sigma}$. To this end, we can rewrite the last three integrands in equation [18] as:

$$\int_0^a \lambda_i(a-\tau,t-\tau) + \lambda_k(a-\tau,t-\tau)d\tau = \sum_0^a 2\bar{\lambda}(a-\tau,t-\tau)\Delta\tau$$

[19]

$$\int_0^a \sum_{k\neq i} \sigma_k d\tau = (N-1)\bar{\sigma}\Delta\tau$$

[20]

$$\int_0^a \sigma_i d\tau = \bar{\sigma}\Delta\tau$$

[21]

where, again, $\Delta\tau$ corresponds to the duration of time acted on by $\bar{\sigma}$.

*5.3. Fitting FOI models to the data*

All versions of the above models (with and without age modification and/or waning multitypic immunity) were fit to the age distribution of cases per year, for each province and the national dataset as a whole using a quasi-Newton (L-BFGS-B) optimization method in the R package 'optim'. The fits of each model to the data were compared via AIC following convergence (Supplementary Table S8). 95% confidence intervals were constructed from the hessian matrix for fits of the FOI, age modification terms, and waning heterotypic immunity.

### Supporting Text 6: Febrile Cohort mNGS, Generation of Consensus DENV Genomes and Phylodynamic and Phylogenetic Analysis

*6.1 Viral sequencing*

Briefly, pathogen mNGS libraries were prepared from isolated pathogen RNA and converted to cDNA Illumina libraries using the NEBNext Ultra II DNA Library Prep Kit (E7645) according to manufacturer's instructions. Library size and concentration were determined using the 4150 Tapestation system, Agilent, and Qubit 4 Flurometer, Invitrogen (for quantitation only). External RNA Controls 103 Consortium collection, ERCC, ThermoFisher, were used as indicators of potential library preparation errors and for input RNA mass calculation. Samples were sequenced on a NovaSeq (Illumina) instrument and an iSeq100 (Illumina) instrument using 150 nucleotide paired-end sequencing. A water ("no template") control was included in each library preparation. All wet lab bench protocols are updated at https://docs.google.com/document/d/1RtNQc1D4or_ys7OxCCBjh4SDIdy7JaI4IE7if8EkHgE/edit.

Raw fastq files were uploaded to the CZID portal, a cloud-based, open-source bioinformatics platform, to identify microbes from metagenomic data (https://czid.org) (24). Potential pathogens were distinguished from commensal flora and contaminating microbial sequences from the environment by establishing a z-score metric based on a background distribution derived from 16 non-templated "water-only" control libraries. Data were normalized to reads mapped per million input reads for each microbe at both species and genus levels. Taxa with z-score less than 1, base pair alignment less than 50 base pairs, NT log(1/e) less than 10 and reads per million (rpM) less than 10 were removed from analysis. Microbial sequences from the samples are available for access in the National Center for Biotechnology Information (NCBI) Sequence Read Archive.

*6.2 Generation of consensus genomes for DENV*

We attempted to construct full-genome DENV sequences from any samples which were confirmed to be DENV-positive by RT-qPCR and which generated at least one reliable contig mapping to any serotype of DENV in the CZID pipeline. To generate full genome consensus sequences, we ran the ARTIC network's Nextflow consensus genome pipeline (25), mapping each sequence to the closest GenBank accession number hit in the original mNGS run of CZID, using a cutoff of 5 reads per nucleotide site to make a consensus call (sites with <5 reads were called as "N"). Sequences were additionally run through the CZID integrated consensus genome pipeline, again mapping to the closest hit identified in GenBank from the original mNGS assembly. Resulting consensus sequences from both assembly pipelines were then aligned with reference sequences and visually examined in Geneious Prime. Raw reads from mNGS were then mapped to each full genome contig in turn and examined manually to determine the correct call for each base pair.

*6.3 Sequence selection for Bayesian timetrees*

For construction of Bayesian timetrees (Fig. 4 CD), we supplemented our own Cambodia sequences with all other Cambodian sequences for DENV-1 and DENV-2 available in GenBank at the time of analysis, selecting all full or partial genome nucleotide sequences >10,000 bp in length up to a collection date of December 31, 2022 (DENV-1: tax id 11053, 197 sequences, including 63 contributed by this study; DENV-2: tax id 11060, 176 sequences, including 120 contributed by this study). We further supplemented these Cambodia sequences with genomes collected from other major Southeast Asian countries (nine), which were Laos, Myanmar, Malaysia, Thailand, Vietnam, Brunei, Indonesia, the Philippines, and Singapore. All countries were represented in both the DENV-1 and DENV-2 datasets. To avoid overrepresenting certain countries outside of Cambodia, we limited sequence selection to a maximum of three randomly selected genomes collected per year from each available year per country, beginning in 2002, the year in which we began our national time series.

*6.4 Sequence selection for Maximum Likelihood phylogeny*

For construction of the maximum likelihood phylogeny visualized in Fig. S18, we included all Cambodian sequences of DENV-1 and DENV-2 available in NCBI, as well as a broadly representative subset of sequences within all known genotypes of DENV-1 (Genotypes I, II, and III) and DENV-2 (Genotypes 1, II, III, IV, V; previously known by their regional names, respectively: American, Cosmopolitan-I/-II/-III, Asian-American, Asian-II, Asian-I) (26).

### Supporting Text 7: The structured, discrete-time epidemic model

To formalize our mechanistic hypotheses of the transmission dynamics underlying the patterns of elevated mean age of dengue infection and expansion of the age distribution of cases witnessed in our data, we developed an age-structured, discrete time, epidemic matrix model to allow us to simulate each hypothetical scenario (27–29).

Under this modeling approach, we can express the population as a vector $\boldsymbol{n}(t)$, divided into both age and epidemic classes. To model the dynamics of four circulating dengue serotypes, we allow for 147 discrete epidemic states and 100 annual age classes. Epidemic states take the following form:

- Susceptible to all serotypes (one state): $S$
- Primary infections (four states): $I_1, I_2, I_3, I_4$
- Recovered from primary infection and protected from reinfection with any serotype by heterotypic immunity (four states): $P_1, P_2, P_3, P_4$
- Protected by monotypic immunity from primary infection but susceptible to other serotypes due to waned heterotypic immunity (four states; here, subscript corresponds to the serotype to which the class is no longer susceptible): $S_1, S_2, S_3, S_4$
- Secondary infections (twelve states): $I_{12}, I_{13}, I_{14}, I_{21}, I_{23}, I_{24}, I_{31}, I_{32}, I_{34}, I_{41}, I_{42}, I_{43}$
- Recovered from secondary infection and protected from reinfection with any serotype by heterotypic immunity (twelve states): $P_{12}, P_{13}, P_{14}, P_{21}, P_{23}, P_{24}, P_{31}, P_{32}, P_{34}, P_{41}, P_{42}, P_{43}$
- Protected by multitypic immunity from secondary infection but susceptible to other serotypes due to waned heterotypic immunity (twelve states; here, subscript corresponds to the serotypes to which the class is no longer susceptible):
  $S_{12}, S_{13}, S_{14}, S_{21}, S_{23}, S_{24}, S_{31}, S_{32}, S_{34}, S_{41}, S_{42}, S_{43}$
- Tertiary infections (24 states):
  $I_{123}, I_{124}, I_{132}, I_{134}, I_{142}, I_{143}, I_{213,}, I_{214}, I_{231}, I_{234}, I_{241}, I_{243,}$
  $I_{312}, I_{314}, I_{321}, I_{324}, I_{341}, I_{342}, I_{412}, I_{413}, I_{421}, I_{423}, I_{431}, I_{432}$
- Recovered from tertiary infection and protected from reinfection with any serotype by heterotypic immunity (24 states):
  $P_{123}, P_{124}, P_{132}, P_{134}, P_{142}, P_{143}, P_{213,}, P_{214}, P_{231}, P_{234}, P_{241}, P_{243,}$
  $P_{312}, P_{314}, P_{321}, P_{324}, P_{341}, P_{342}, P_{412}, P_{413}, P_{421}, P_{423}, P_{431}, P_{432}$
- Protected by multitypic immunity from secondary infection but susceptible to other serotypes due to waned heterotypic immunity (24 states; here, subscript corresponds to the serotypes to which the class is no longer susceptible):
  $S_{123}, S_{124}, S_{132}, S_{134}, S_{142}, S_{143}, S_{213,}, S_{214}, S_{231}, S_{234}, S_{241}, S_{243,}$
  $S_{312}, S_{314}, S_{321}, S_{324}, S_{341}, S_{342}, S_{412}, S_{413}, S_{421}, S_{423}, S_{431}, S_{432}$
- Quaternary infections (24 states):
  $I_{1234}, I_{1243}, I_{1324}, I_{1342}, I_{1423}, I_{1432}, I_{2134,}, I_{2143}, I_{2314}, I_{2341}, I_{2413}, I_{2431,}$
  $I_{3124}, I_{3142}, I_{3214}, I_{3241}, I_{3412}, I_{3421}, I_{4123}, I_{4132}, I_{4213}, I_{4231}, I_{431}2, I_{4321}$
- Temporary heterotypic immunity to all serotypes (one state): $P_{ms}$
- Longterm multitypic immunity to all serotypes (one state): $P_m$

From above, the population vector takes the following form:

$$\boldsymbol{n}(t) = \left(S_{1,t,} \dots S_{a_{max},t}, I_{1_{1,t}} \dots I_{1_{a_{max},t}}, I_{2_{1,t}} \dots I_{2_{a_{max},t}}, \dots \dots \dots \dots \dots \dots \dots \dots \; P_{m_{1,t}} \dots P_{m_{a_{max},t}}\right)$$

where $a_{max}$ indicates the oldest age class in the model (here, 100 years).

Next, we investigated epidemic and demographic transitions for this population using a transition matrix. Following (29), we initially ignore demographic transitions (survival and aging), to describe transitions between the above 147 epidemic states within a single age class $a$ according to:

$$\begin{pmatrix} 1-4\bar{\lambda} & 0 & 0 & 0 & 0 & 0 & 0 & 0 & 0 & 0 & \dots \\ \bar{\lambda} & 1-r & 0 & 0 & 0 & 0 & 0 & 0 & 0 & 0 & \dots \\ \bar{\lambda} & 0 & 1-r & 0 & 0 & 0 & 0 & 0 & 0 & 0 & \dots \\ \bar{\lambda} & 0 & 0 & 1-r & 0 & 0 & 0 & 0 & 0 & 0 & \dots \\ \bar{\lambda} & 0 & 0 & 0 & 1-r & 0 & 0 & 0 & 0 & 0 & \dots \\ 0 & r & 0 & 0 & 0 & 1-\omega & 0 & 0 & 0 & 0 & \dots \\ 0 & 0 & r & 0 & 0 & 0 & 1-\omega & 0 & 0 & 0 & \dots \\ 0 & 0 & 0 & r & 0 & 0 & 0 & 1-\omega & 0 & 0 & \dots \\ 0 & 0 & 0 & 0 & r & 0 & 0 & 0 & 1-\omega & 0 & \dots \\ 0 & 0 & 0 & 0 & 0 & \omega & 0 & 0 & 0 & 1-3\bar{\lambda} & \dots \\ \dots & \dots & \dots & \dots & \dots & \dots & \dots & \dots & \dots & \dots & \dots \\ 0 & 0 & 0 & 0 & 0 & 0 & 0 & 0 & 0 & 0 & 1 \end{pmatrix} \quad [20]$$

where $\bar{\lambda}$ corresponds to the mean probability of transmission for a single serotype, $r$ is the probability of recovery (equal to 1 if the model is simulated in biweekly timesteps according to the generation time of dengue), and $\omega$ is the probability of waning heterotypic immunity. The transition pattern repeats itself across the full iteration of the matrix, with the number of serotypes moving out of each susceptible class (by N*$\bar{\lambda}$) decreasing from 4 in the first susceptible class to one in the last susceptible class preceding quaternary infections. Please see our open access github repository (https://github.com/brooklabteam/cambodia-dengue-national) for the full matrix specification.

The above transition matrix describes the epidemic transitions assuming four endemic circulating serotypes in the system. This matrix can be modified to account for only three strains (e.g. some of the $\bar{\lambda}$ values become 0 and expressions of $1-3(\bar{\lambda})$ become $1-2(\bar{\lambda})$ instead) or to account for differing dynamical assumptions.

For simplicity, we first explored the dynamics of three endemic circulating strains, then investigated hypotheses of genotype invasion and replacement, modeling the new genotype as if equivalent to a new serotype which could reinfect all prior exposures but specifically replaced a single exposure genotype. We also considered alternative hypotheses of increasing detectability of tertiary exposures under the endemic-three serotype simulation. Results would be qualitatively similar if instead simulating enhanced detections of both tertiary and quaternary infections under an endemic four-serotype simulation.

From above, again following (29), we next add in demography, to construct the full transition matrix $\boldsymbol{A}(\boldsymbol{n}(t))$, which we use to project the entire population forwards (via aging, mortality and epidemic transitions) according to:

$$\boldsymbol{A}(\boldsymbol{n}(t)) = \begin{pmatrix} s_1(1-u_1)\boldsymbol{A}_1 & 0 & 0 & 0 & 0 & 0 \\ s_1u_1\boldsymbol{A}_1 & s_2(1-u_2)\boldsymbol{A}_2 & 0 & 0 & 0 & 0 \\ 0 & s_2u_2\boldsymbol{A}_2 & s_3(1-u_3)\boldsymbol{A}_3 & 0 & 0 & 0 \\ 0 & 0 & s_3u_3\boldsymbol{A}_3 & 0 & 0 & 0 \\ \dots & \dots & \dots & \dots & \dots & 0 \\ 0 & 0 & 0 & 0 & 0 & .s_z\boldsymbol{A}_z \end{pmatrix} \quad [21]$$

where $s_a$ is the probability that an individual of age class $a$ survives to the next timestep, $u_a$ is the probability of aging out of age class $a$ and $\boldsymbol{A}_1$, $\boldsymbol{A}_2$, etc. correspond to the epidemic transition matrix in equation [20].

We then project the dynamics of the population as a whole forward by multiplying the population vector by the transition matrix and adding in a vector of annual births:

$$\boldsymbol{n}(t+1) = \boldsymbol{A}(\boldsymbol{n}(t))\boldsymbol{n}(t) + \boldsymbol{B}(t)$$

[22]

where $\boldsymbol{B}(t)$ is a vector of the number of births at time $t$:

$$\boldsymbol{B}(t) = (\boldsymbol{B}(t), 0, 0, \dots 0)^T$$

[23]

From above, we simulated the dynamics of the four hypotheses outlined in the main text (H0: standard demography; H1: increasing tertiary case detection through time; H2: novel genotype invasion with clade replacement and waning immunity within the serotype in 2019 and in 2007; H3: novel genotype invasion with clade replacement but standard immunity; and H4: novel genotype invasion with clade replacement but increasing tertiary case detection through time. For each simulation, we tracked infection status per age category through time.

We simulated in biweekly timesteps, according to the generation time of the pathogen. We initiated our simulations with an initial population vector at $t$=0 using the proportional age distribution of Cambodia from 1950, as recovered from (30) and 5 infected individuals in the first age class for each serotype under circulation. We first simulated dynamics out to equilibrium for 20 years, then, in the last 22 years of the time series, we introduced parameters that aimed to approximate the real-world dynamics of dengue in Cambodia over the past two decades.

Parameters were fixed at the following rates, which we converted to biweek probabilities prior to input into the transition matrix in equation [20]:

- $\bar{\lambda}$, the single-serotype FOI, was fixed at values estimated at the national level via the fitting of Ferguson-Cummings catalytic model in Fig. 3, from 1999-2020.
    - $\bar{\lambda}$ was further modified by age according to parameters estimated for the first half of the time series (2002-2010) in Fig. 3 analyses.
    - We additionally introduced intra-annual climate variation to the FOI, by averaging biweekly transmission rates as estimated for TSIR across all provinces and years and rescaling them from 0.5 to 1.5 to allow for subtle seasonal dampening or amplification of the FOI.
- Births ($\boldsymbol{B}(t)$), were input annually outside of the transition matrix as shown in equation [22], corresponding to publicly available birth rates through time reported by the World Bank (11).
- Age-specific death rates (from which we calculated biweekly survival rates per age class) were obtained from the United Nations databank (31).
- Infected individuals were assumed to recover from infection in a single biweekly timestep (rate $r$).
- Heterotypic immunity was assumed to wane ($\omega$) at a rate of $\frac{1}{2}$ yrs$^{-1}$, as estimated in the literature (32).
- Waning homotypic immunity was not explicity modeled but was accounted for by allowing for the introduction of a fourth infection to an endemic, three-serotype model at a specified timepoint. In the case of these introductions, we modeled genotype replacement by silencing the FOI for the original strain following the introduction of the second strain within a serotype, then explored differences in detectability for diverse immunological states, as outlined in the main text.

Antibody-dependent enhancement was not considered in this model.

After simulations of hypotheses were complete, we finally fit our Ferguson-Cummings catalytic model from Fig. 3 to the resulting distribution of cases by age, by year, to attempt to recover the input FOI across the time series. We additionally fit a parameter of waning immunity ($\sigma$) to each simulated dataset after estimating and fixing FOI, to evaluate the conditions under which this model was improved by consideration of waning immunity.

All code and data needed to reproduce all analyses and run all simulations is publicly available in our GitHub repository: https://github.com/brooklabteam/cambodia-dengue-national

### Supplementary Figures

**Figure S1.** Annual biweekly mean temperatures ($^0$C) for Cambodia, aggregated by province and year.

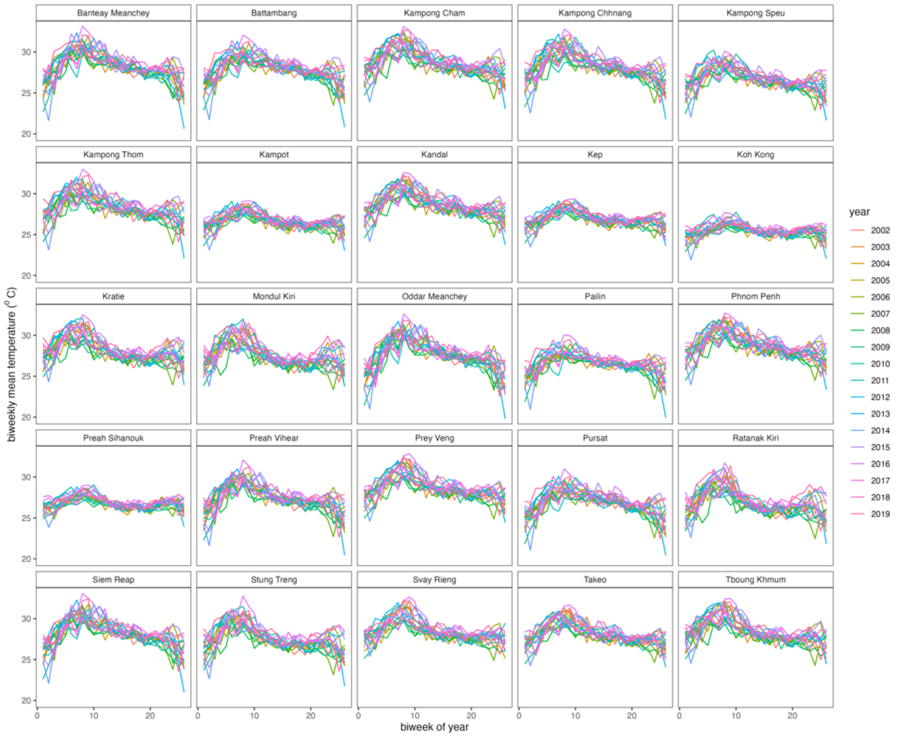


**Figure S2.** Annual biweekly total precipitation (mm) for Cambodia, aggregated by province and year.

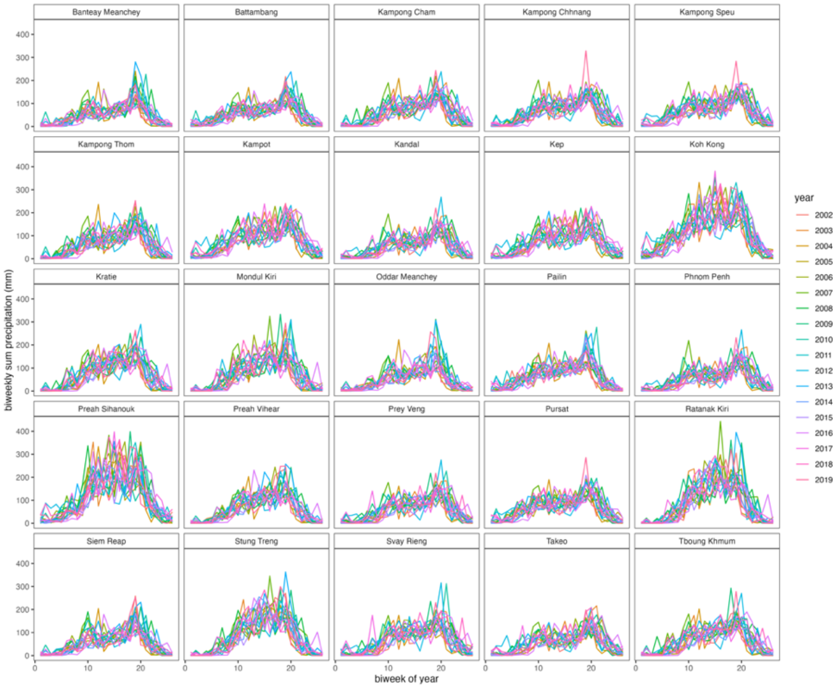

**Figure S3.** Interannual trends in mean biweekly temperature ($^0$C) for Cambodia, aggregated by province. Black lines give GAM predictions of interannual trends, with 95% confidence intervals by standard error shown as narrow shading around mean projection (Table S1).

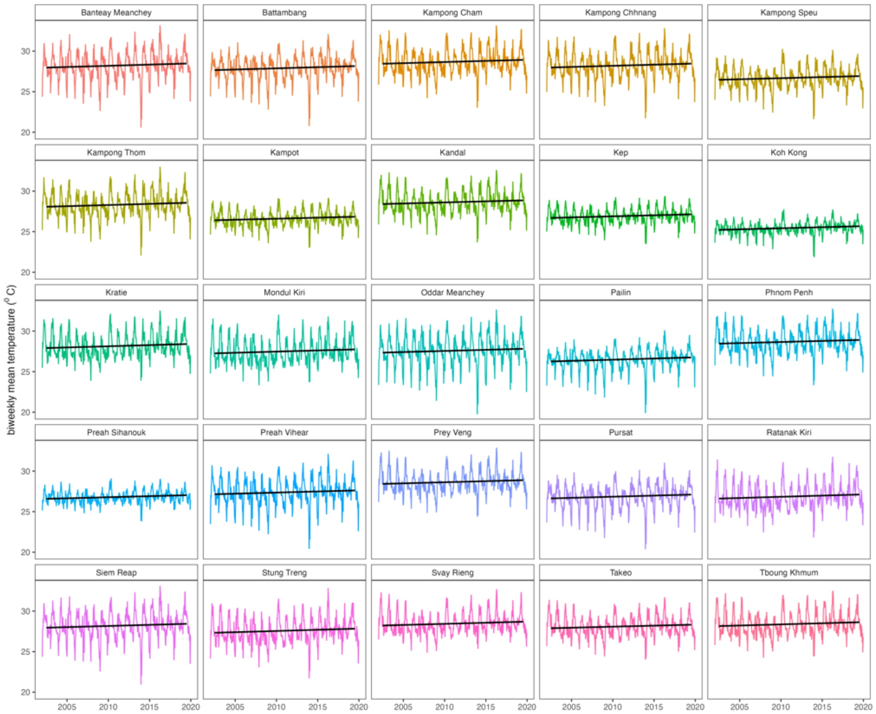


**Figure S4.** Interannual trends in total mean precipitation (mm) for Cambodia, aggregated by province. Black lines give GAM predictions of interannual trends, with 95% confidence intervals by standard error shown as narrow shading around mean projection (Table S1).

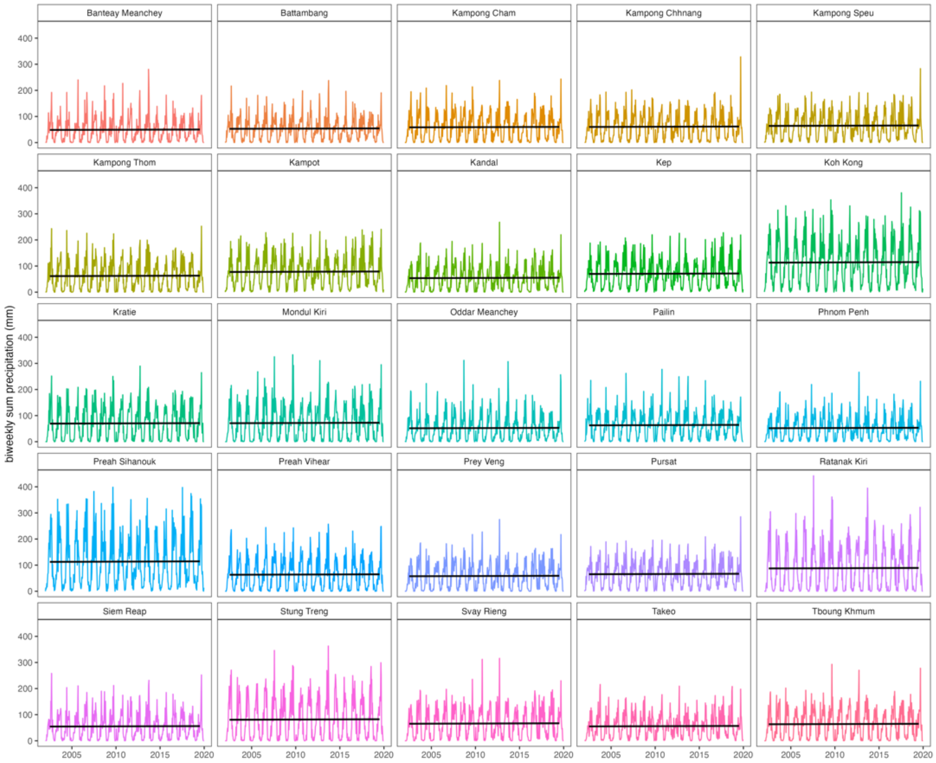

**Figure S5.** Identifying anomalous years in the temperature time series for Cambodia. **A** Partial effect of year, as a factor input as a smoothing spline, on response variable of mean biweekly temperature per province from GAMs (Table S1). Anomalously hot and cool years for which confidence intervals on effect size do not cross zero are colored, respectively, red and blue. **B** Map of Cambodia with provinces indicated by color. **C** Z-scores of temperature time series, with epidemic years indicated by vertical lines. Provinces are arranged by latitude of centroid, from south to north and colored according to map**.** The top panel gives the distribution of annual temperature z-scores across all provinces.

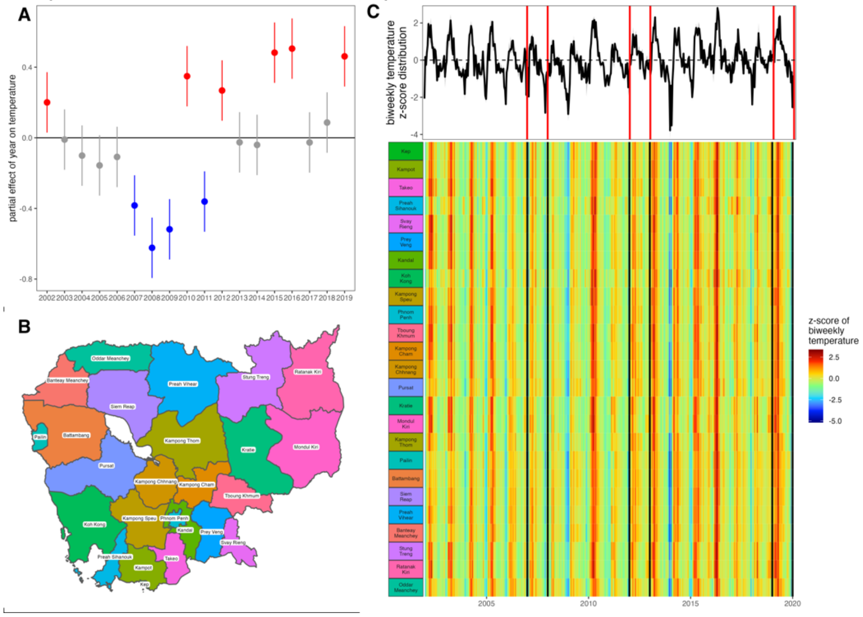


**Figure S6.** Identifying anomalous years in the precipitation time series for Cambodia. **A** Partial effect of year, as a factor input as a smoothing spline, on response variable of total biweekly precipitation per province from GAMs (Table S1). Anomalously wet and dry years for which confidence intervals on effect size do not cross zero are colored, respectively, red and blue. **B** Map of Cambodia with provinces indicated by color. **C** Z-scores of precipitation time series, with epidemic years indicated by vertical lines. Provinces are arranged by latitude of centroid, from south to north and colored according to map**.** The top panel gives the distribution of annual precipitation z-scores across all provinces.

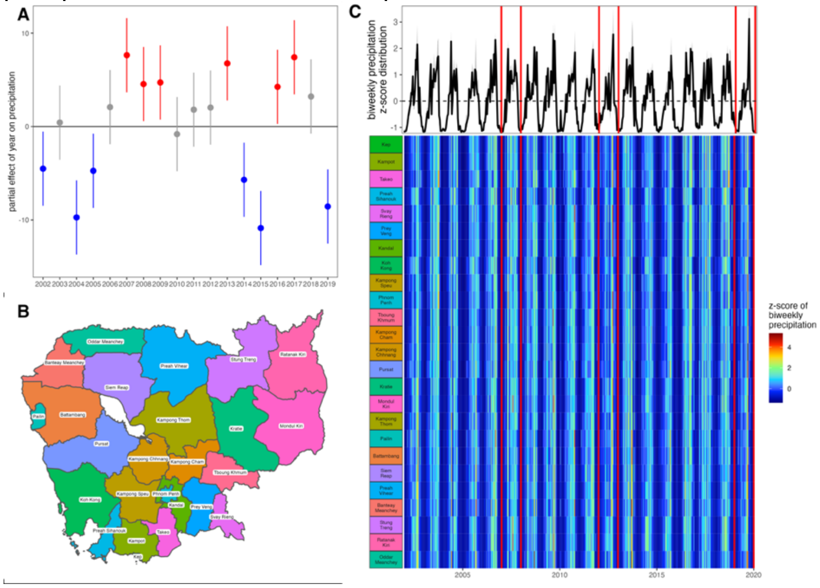

**Figure S7.** Impact of climate predictors on biweekly transmission rate ($\beta$) from TSIR, 2002-2006. **A** Coefficients from linear mixed effects regression, **B** coefficients from linear mixed effects regression for precipitation paired with Brière function transformation for temperature, and **C** coefficients from generalized additive model, fitted to climate variable associations with transmission rates for the 2002-2006 interepidemic period (Table S4).

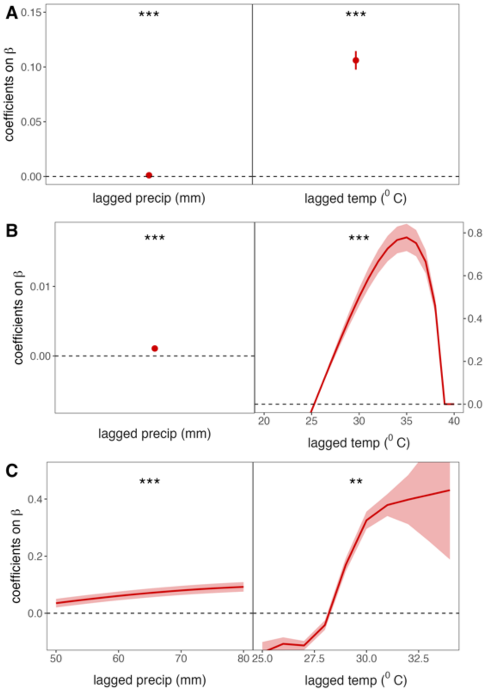


**Figure S8.** Impact of climate predictors on biweekly transmission rate ($\beta$) from TSIR, 2008-2011. **A** Coefficients from linear mixed effects regression, **B** coefficients from linear mixed effects regression for precipitation paired with Brière function transformation for temperature, and **C** coefficients from generalized additive model, fitted to climate variable associations with transmission rates for the 2008-2011 interepidemic period (Table S4).

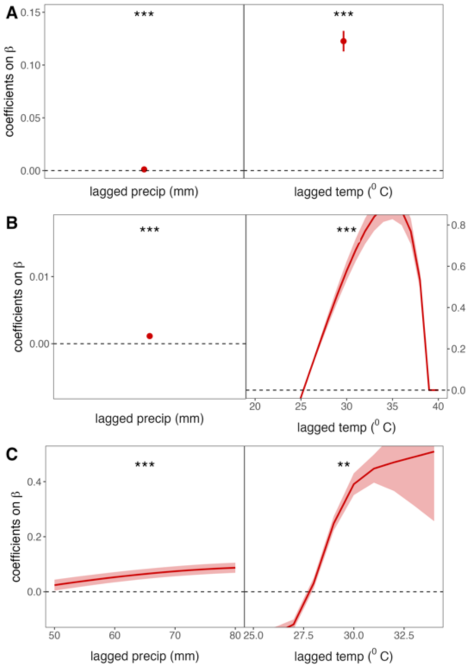

**Figure S9.** Impact of climate predictors on biweekly transmission rate ($\beta$) from TSIR, 2013-2018. **A** Coefficients from linear mixed effects regression, **B** coefficients from linear mixed effects regression for precipitation paired with Brière function transformation for temperature, and **C** coefficients from generalized additive model, fitted to climate variable associations with transmission rates for the 2013-2018 interepidemic period (Table S4).

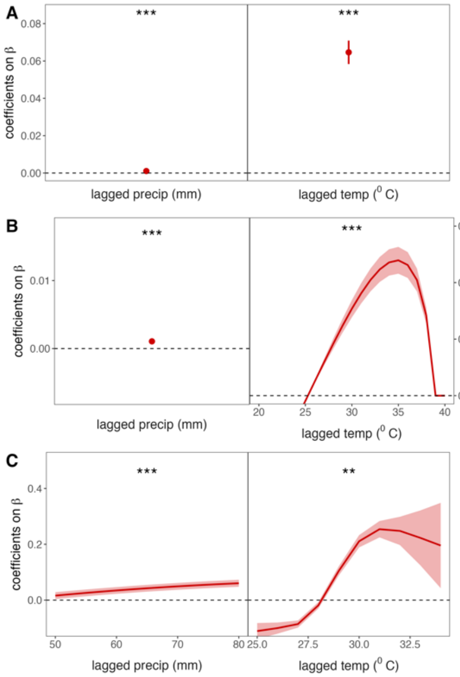


**Figure S10.** Biweekly dengue transmission rates, by province, from TSIR. Province-specific transmission rates ($\beta$) are colored by inter-epidemic period, according to legend, with TSIR-fitted $\beta$ shown as a solid line and climate-projected $\beta$ from epidemic year regressions shown as a dashed line. The 95% confidence intervals by standard error are shown in translucent shading around the mean $\beta$ estimates.

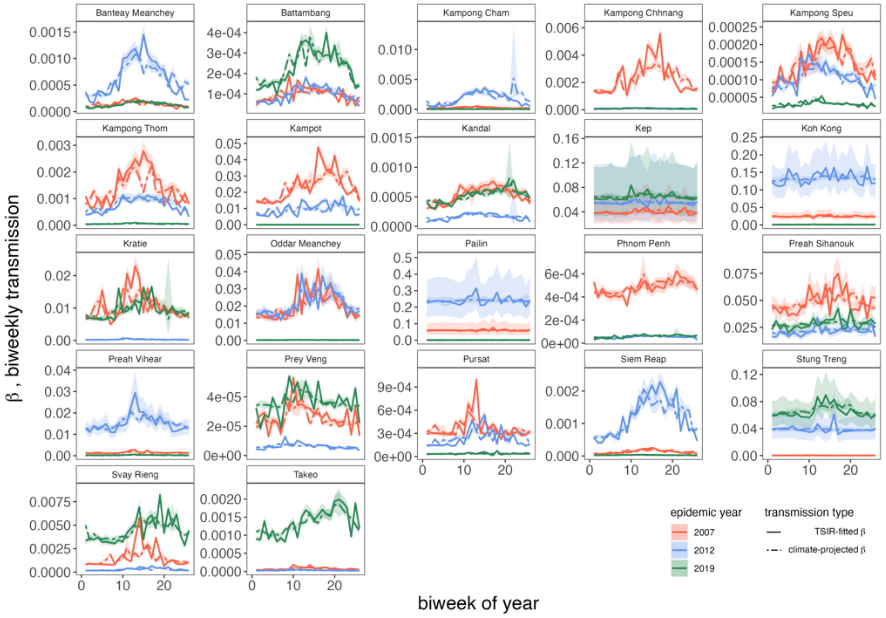

**Figure S11.** Average wavelet power for raw dengue incidence time series over **A** annual and **B** multiannual time horizons. **C** Proportion of provinces with which a focal province demonstrates statistically significant cross-wavelet power in the annual incidence time series or **D** between reconstructed cycles over a 5-year time horizon. Top panels give the biweekly distribution of wavelet power at annual (**A**) or multiannual (**B**) scales or the range in proportion of provinces exhibiting significant cross-wavelet power at annual (**C**) or multiannual (**D**) scales. Provinces are arranged by latitude of centroid, from south to north, colored after map in Fig. S5-S6.

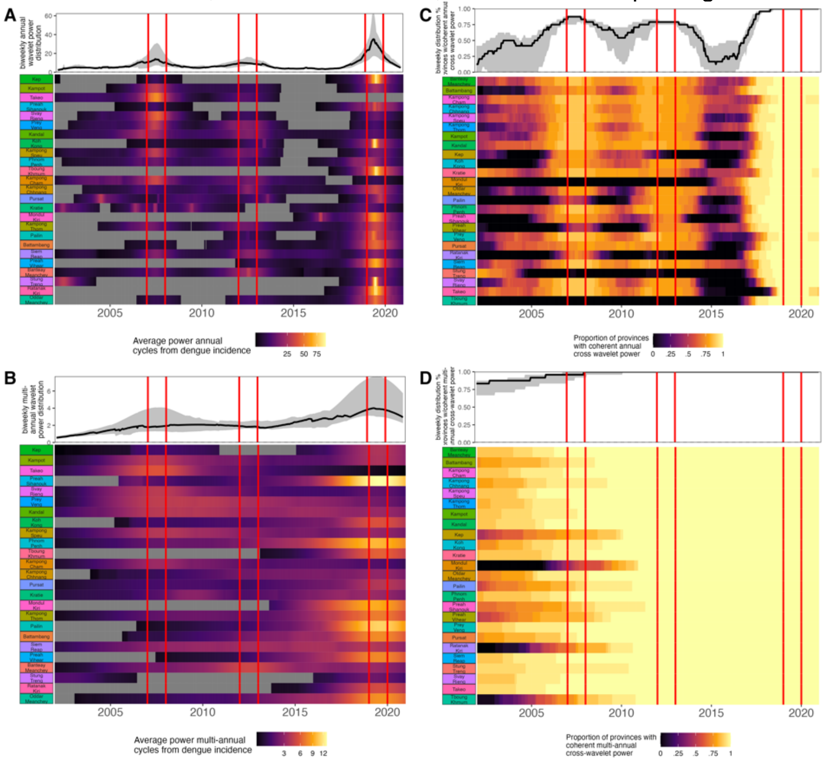


**Figure S12.** Predictors of annual synchronicity in annual dengue incidence between provinces. **A** Coefficient of the fixed interaction of province and geographic distance on the Pearson's correlation coefficient ($\rho$). **B** Partial effect of year (input as a factor), **C** mean biweekly temperature of focal province, **D** mean total annual precipitation of focal province, and **D** mean population size across the time series of focal province on $\rho$. Predictors with significant positive slopes are colored red, predictors with significant negative slopes colored blue, and insignificant predictors shaded gray (Table S6).

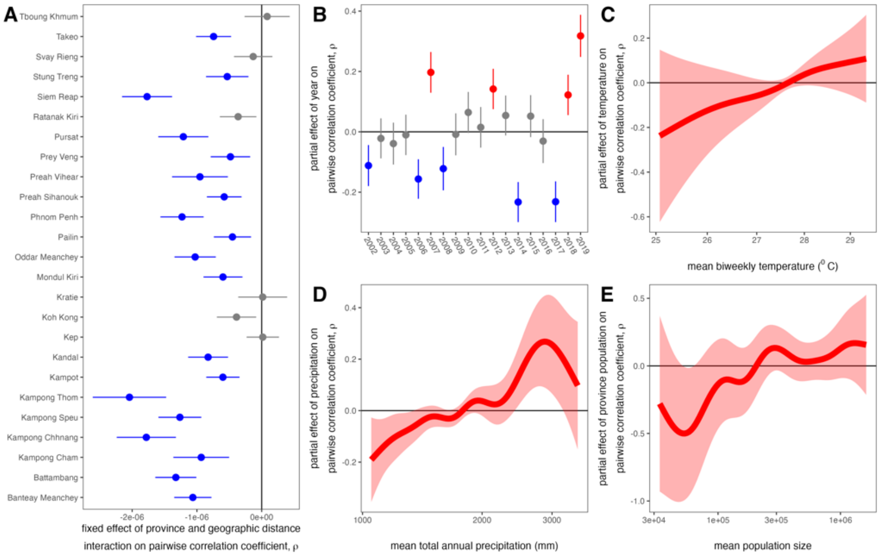

**Figure S13.** Cross-wavelet power between dengue dynamics and climate variables. Average cross wavelet power between biweekly raw dengue incidence and biweekly time series of **A** mean temperature and **B** total precipitation by province. Average cross-wavelet power between biweekly reconstructed multiannual dengue cycles and biweekly time series of **C** mean temperature and **D** total precipitation by province over a 5-year time horizon. **E** Average cross-wavelet power between monthly reconstructed multiannual dengue cycles and ONI by province. Top panels give the range of observed mean cross-wavelet power across all provinces. Provinces are arranged by latitude of centroid, from south to north, colored after map in Fig. S5-S6.

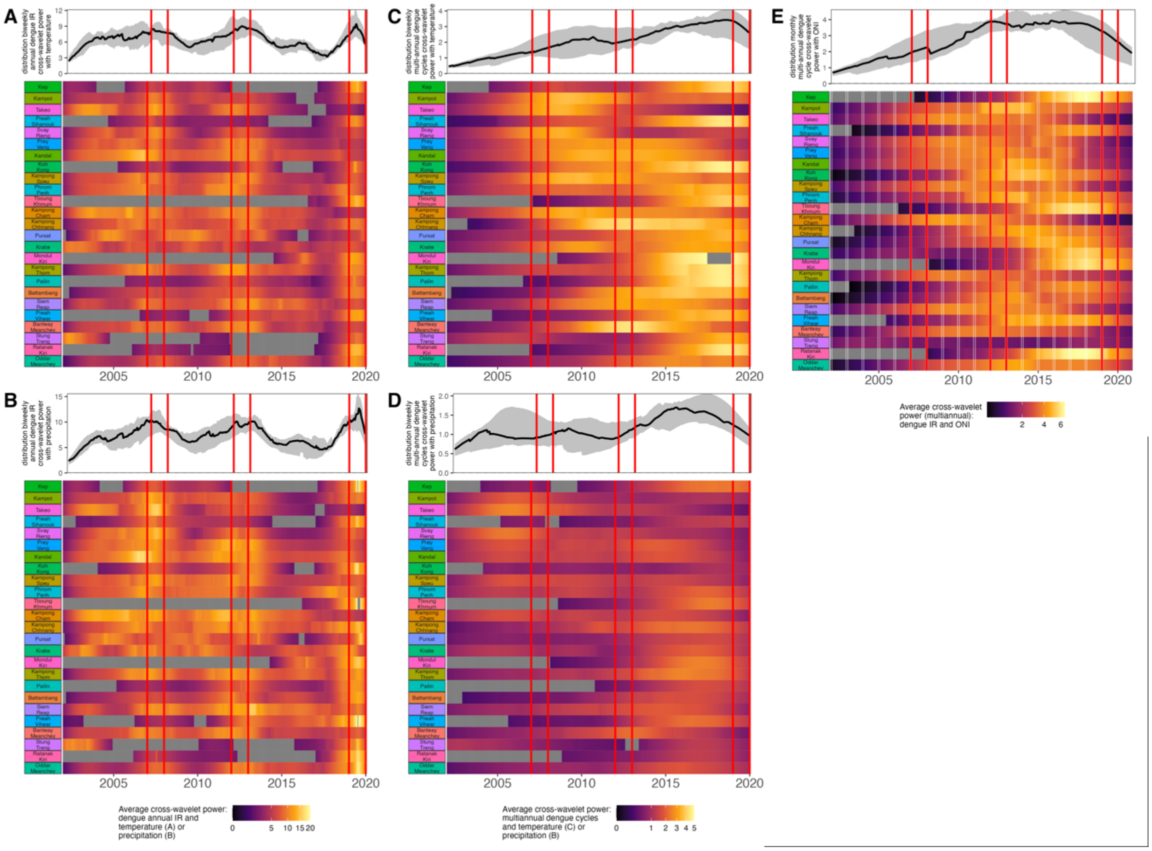


**Figure S14. A** Publicly available demographic data on births, deaths, and total population size preceding and overlapping the NDCP dengue time series. **B** Mean period duration of reconstructed multiannual dengue cycles for Cambodia at both national (black line) and province (colored corresponding to legend) levels across the duration of the NDCP dataset (2002-2020).

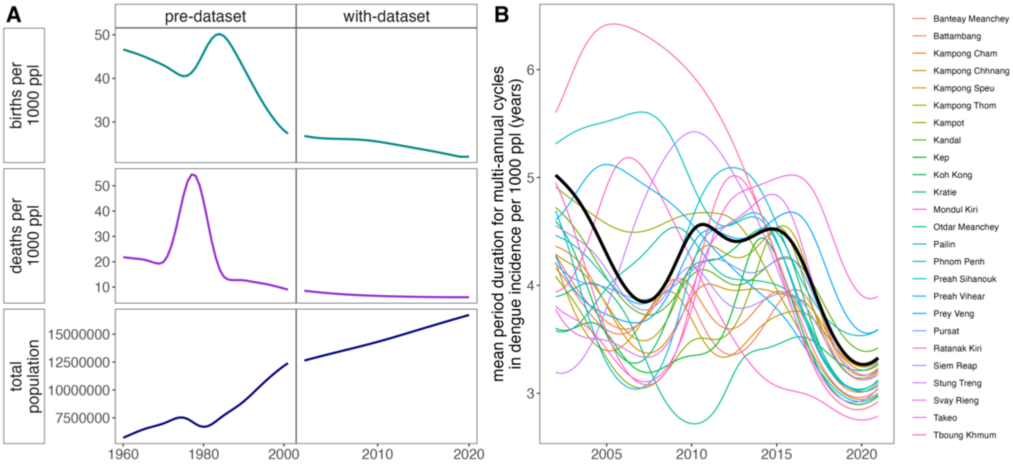

**Figure S15.** Age distribution of reported cases by province, with violin plots highlighting changes in the interquartile range by year. The interannual trend in the mean age of dengue infection per province is plotted as a solid black line across each province subplot, with 95% confidence intervals by standard error shown as a narrow, translucent band behind it (Table S7). Epidemic years (2007, 2012, 2019) are highlighted by dashed lines in the background.

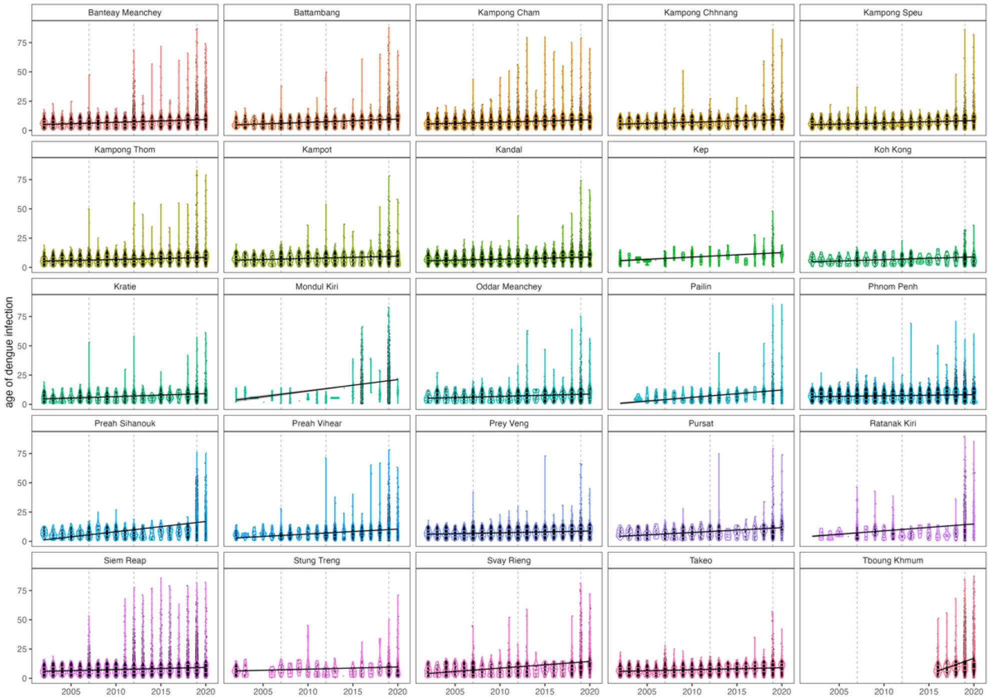


**Figure S16.** Annual estimates for the force of infection by year for 24 of 25 Cambodian provinces and the national time series (Tboung Khmum excluded). FOI was estimated from the birth year of the oldest individual per province, as shown collectively in Fig. 3C, using the Ferguson-Cummings catalytic model, assuming reported cases to represent secondary infections (19, 21, 22). 95% confidence intervals from the hessian matrix are shown as translucent shading.

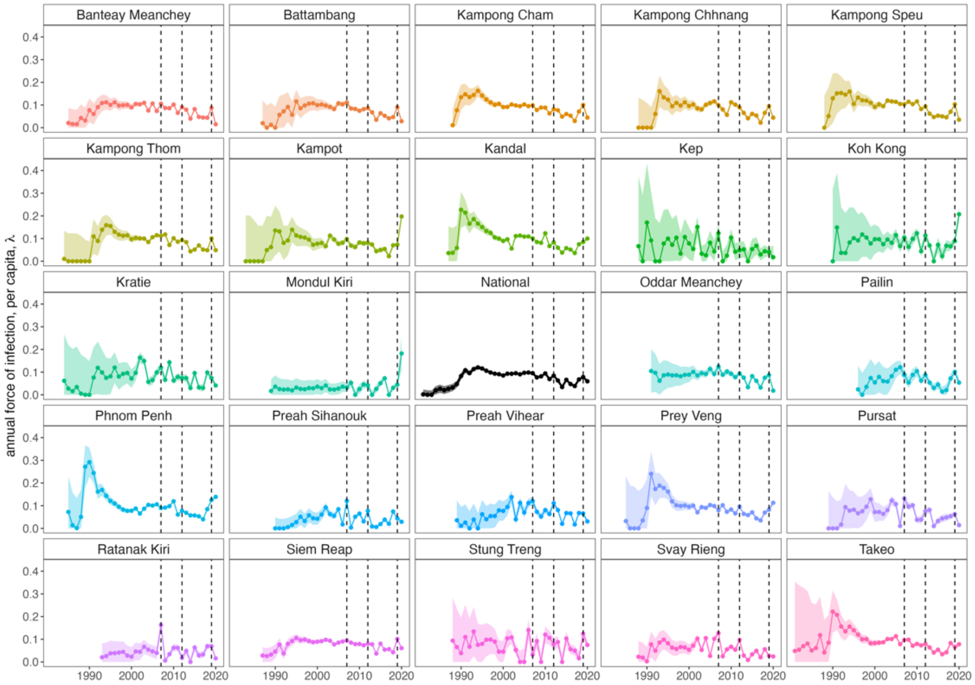

**Figure S17.** Cumulative proportion of cases by age by year by province, with data shown as dotted lines and model projections using FOI estimates from Fig. S16, in addition to age modification of the FOI and a rate of waning multitypic immunity in 2019 and 2020, as shown in Fig. 3D-E of the main text.

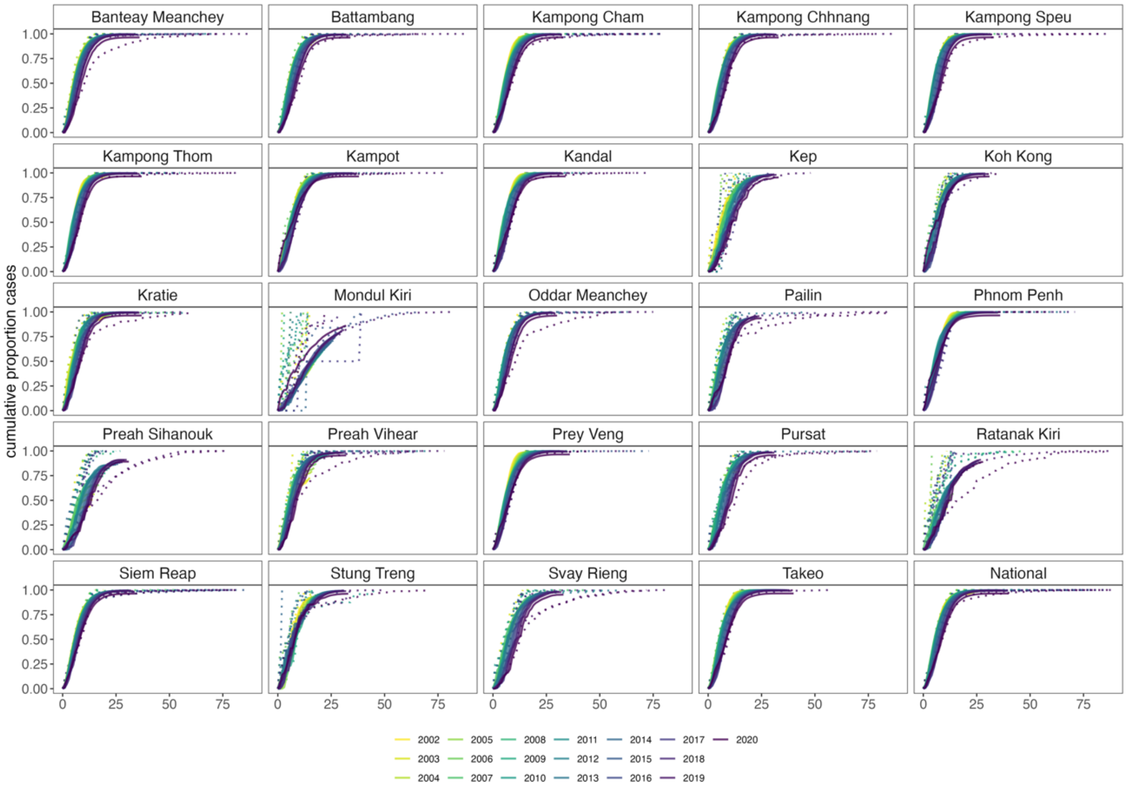

**Fig. S18.** Maximum likelihood phylogenetic tree constructed in RAxML (33) to illustrate how newly-contributed **A** DENV-1 and **B** DENV-2 sequences relate to all known genotypes of the corresponding serotype. Tips are colored by genotype within each serotype, with our Cambodia sequences highlighted in pink and historical Cambodia sequences available in GenBank depicted in purple. Clade bars highlight the extent of each genotype. Trees were constructed using a GTR+I+G4 nucleotide substitution model and rooted in DENV-4 (accession number NC_002640).

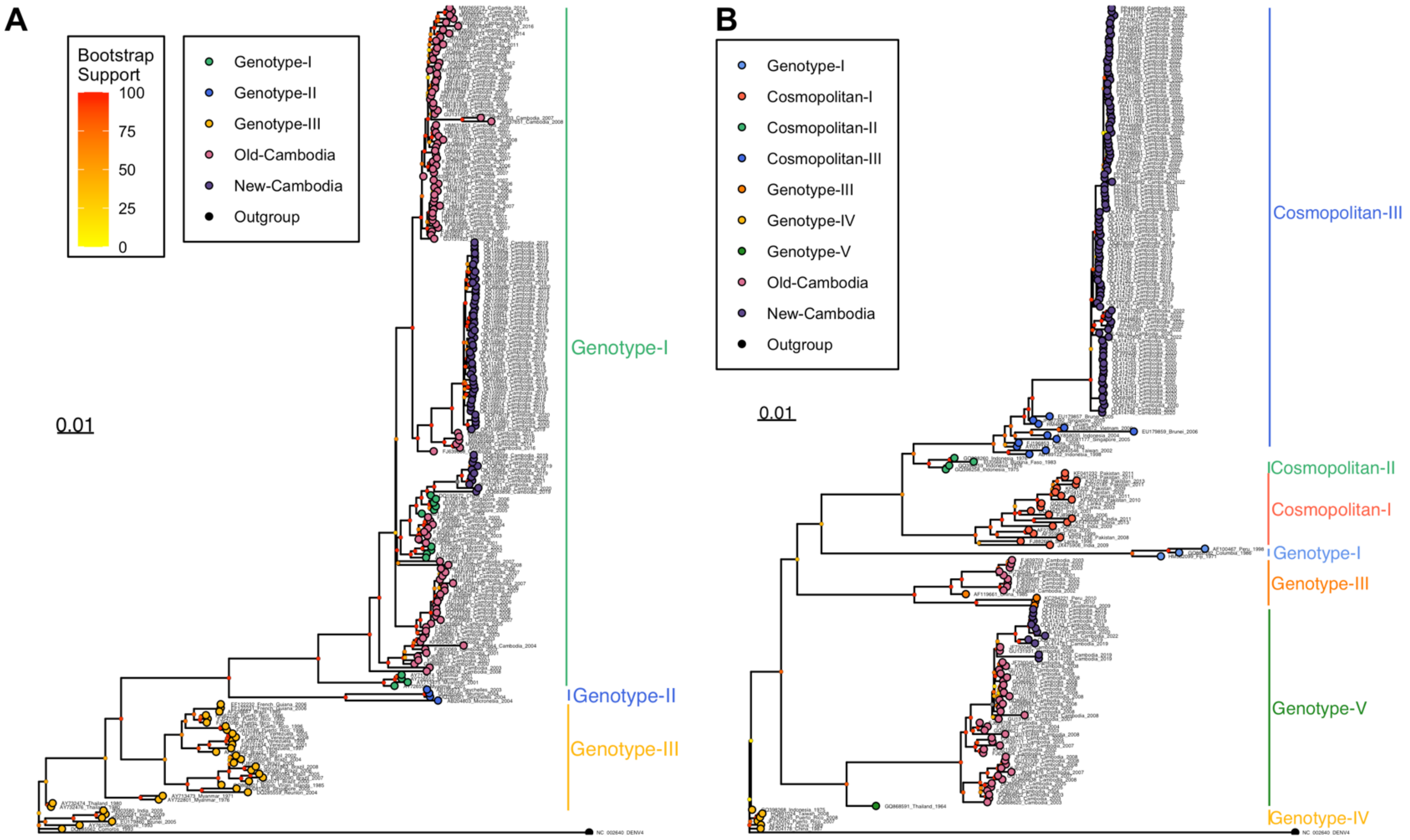

**Figure S19.** Geographic structuring of evolutionary relationships for DENV within Cambodia. **A** Map of Cambodia highlighting location of DENV-1 and **B** DENV-2 full genome sequences contributed to NCBI from our study, with sequences grouped and colored within 10 km radii. **C** DENV-1 and **D** DENV-2 phylogeny subsets from Fig. 4 Bayesian phylogenies in the main text. Cambodia tips are colored corresponding to geolocation of collection, corresponding to panels B and C. Nodes with >0.9 posterior support are colored black and nodes with <0.9 support are colored white.

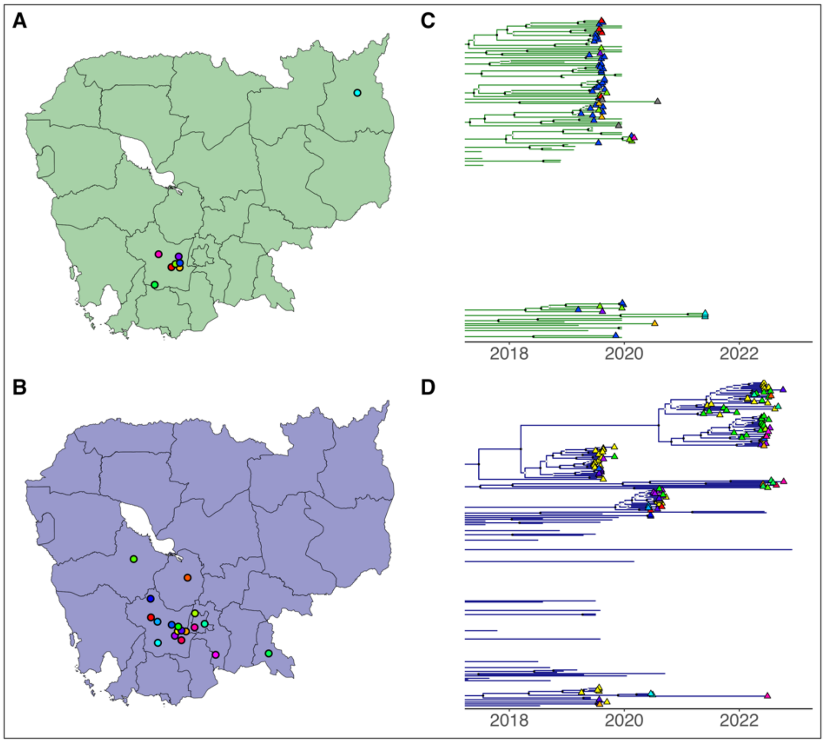


**Figure S20.** Proportion of geolocated sequence pairs from panel A of Fig. 4 (main text) for DENV-1 (*green*) and DENV-2 (*blue*) genomes derived from the same transmission chain across progressively longer Euclidean distances. Here, we explore alternative thresholds in tMRCA for the criteria by which sequences are classed as belonging to the same transmission chain, as indicated by color shading shown in legend.

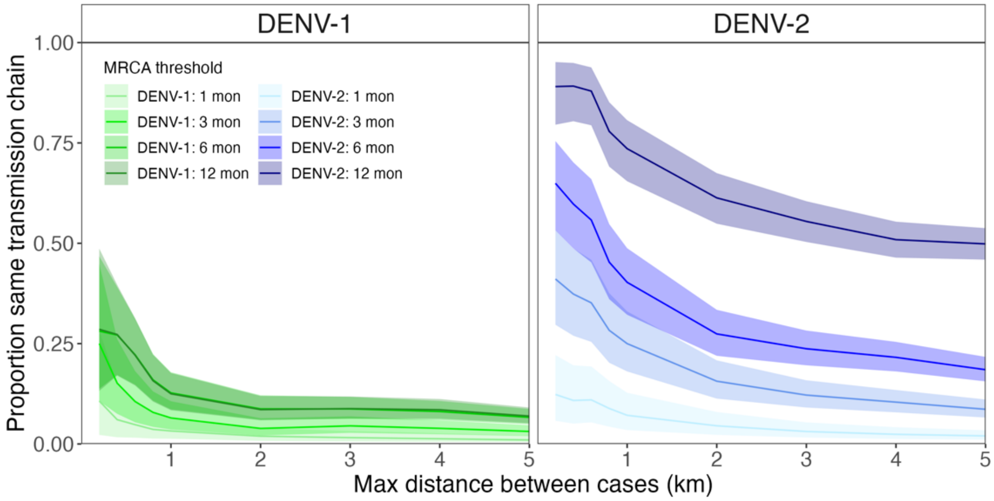

**Fig S21.** Alternative simulations of hypothetical drivers of the age distribution of cases as shown in the main text (Fig. 5). Both simulations shown here model a 2019 introduction of a novel genotype that replaces a prior genotype in an endemic three-serotype system. H3 assumes no change in assumptions regarding immunity and allows for detection of secondary infections only. H4 assumes monotonically increasing tertiary case detectability through time, from 0-15% detectability across the time series. Panel **A** shows the total observed case counts (solid line = mean FOI; translucent shading = 95% confidence interval for FOI), **B** the age distribution of cases by year (secondary = black; tertiary = blue), and **C** the cumulative proportion of cases by age through time under each hypothesis. Under these detection assumptions, recovered data in panels **A** and **B** are not distinguishable from, respectively, H0 demographic simulations or H1 simulations of increasing tertiary case detection in the absence of genotype introduction, as presented in the main text.

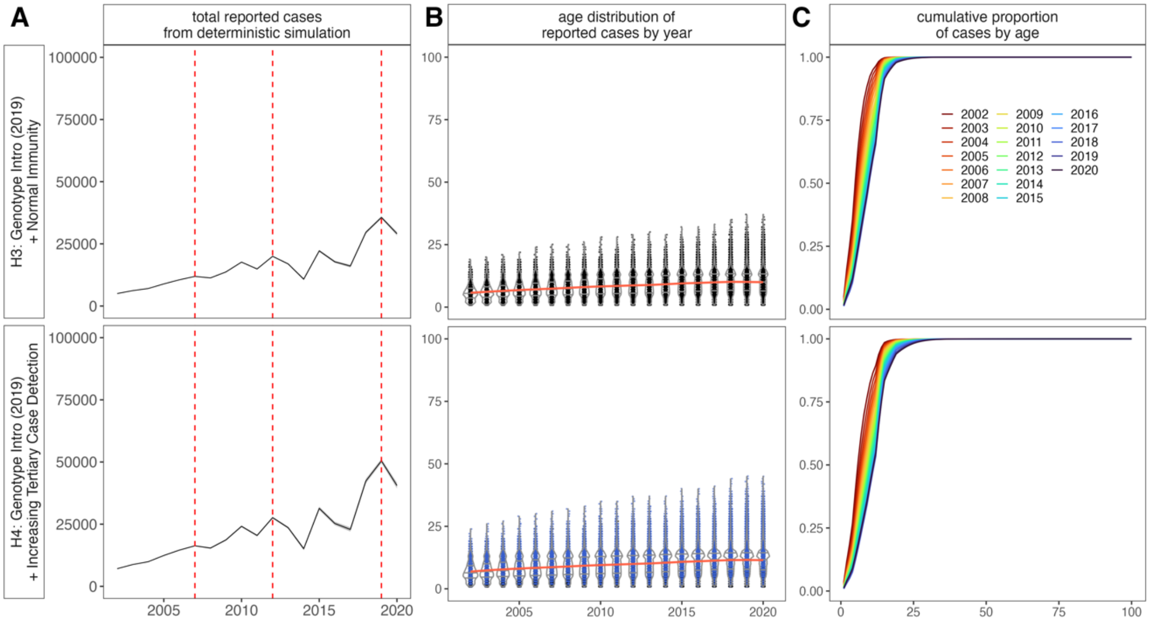


**Figure S22. A** Force of infection and **B** rate of waning multitypic immunity estimates recovered from fitting of Ferguson-Cummings catalytic model to the age-structured time series of cases for our simulated epidemic hypotheses from Fig. 5 (main text). 95% confidence intervals from the hessian matrix are shown as translucent shading. Actual input FOI is shown in **A** as a black line. As expected, only hypothesis H2 2019 (genotype replacement + waning immunity) recovered any signal of waning multitypic immunity, as witnessed in the data.

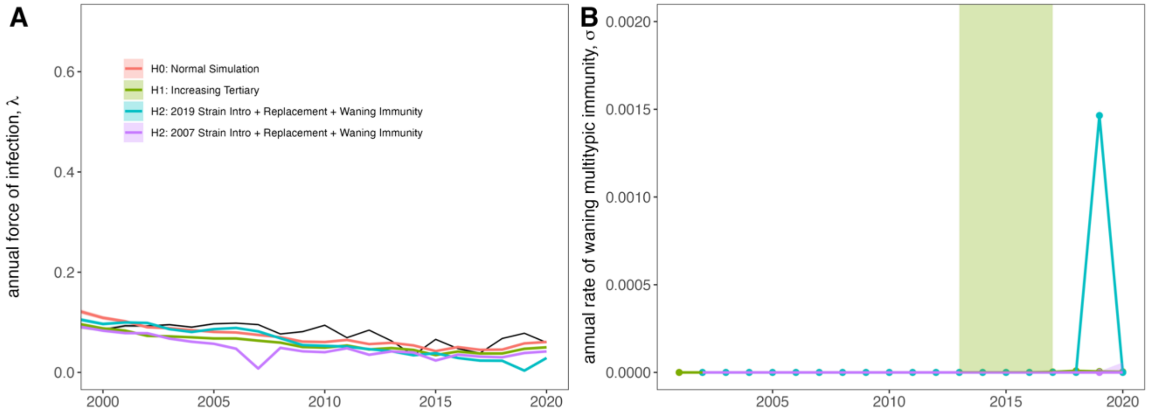

**Supplementary Tables**

*All supplementary tables have been stored in Supplementary Dataset 1, accompanying the main text of the paper. We here provide a brief bibliography of the tables listed:*

| Table Number | Table Title |
|---|---|
| Table S1 | Generalized additive model summaries for climate data |
| Table S2 | Fitted TSIR parameters by province |
| Table S3 | Optimal climate lags to predict biweekly transmission, by inter-epidemic period and province |
| Table S4 | Summary of regression models relating lagged temperature and precipitation to transmission |
| Table S5 | Fraction of increased susceptibles needed to recapture epidemic year caseloads, with and without climate-informed transmission rate |
| Table S6 | Generalized additive model summary for predictors of synchronicity |
| Table S7 | Generalized additive model summary for age of dengue infection |
| Table S8 | FOI fits and model selection with age modifiers and waning multitypic immunity |
| Table S9 | GenBank accession numbers of DENV genomes added in part with this study |
| Table S10 | Linear regression fits for mean age of dengue infection in genotyped cases |

### SI References


1. Copernicus Climate Change Service Climate Data Store (CDS): Copernicus Climate Change Service (C3S), ERA5: Fifth generation of ECMWF atmospheric reanalyses of the global climate. Deposited 2017.

2. AmeriGEOSS Community Platform DataHub., Cambodia Admin Boundaries Level 1. Deposited 2023.

3. J. E. Manning, *et al.*, Development of inapparent dengue associated with increased antibody levels to *Aedes aegypti* salivary proteins: A longitudinal dengue cohort in Cambodia. *The Journal of Infectious Diseases* (2021). https://doi.org/10.1093/infdis/jiab541.

4. S. N. Wood, mgcv: GAMs and Generalized Ridge Regression for R. *R News* **1/2**, 20–24 (2001).

5. W. O. Kermack, A. G. McKendrick, Contributions to the mathematical theory of epidemics II: The problem of endemicity. *Proceedings of the Royal Society A* **115**, 55–83 (1932).

6. R. M. Anderson, R. M. May, *Infectious Diseases of Humans: Dynamics and Control* (Oxford University Press, 1991).

19. Hugo. Muench, *Catalytic models in epidemiology.* (Harvard University Press, 1959).
